# Human inherited RORγT deficiency encompasses genetic heterogeneity, T cell deficiency, and clinical homogeneity

**DOI:** 10.64898/2026.07.18.26358075

**Authors:** Iris Fagniez, Miyuki Tsumura, Antoine Guerin, Hassan Abolhassani, Samin Sharafian, Mehrnaz Mesdaghi, Toyoki Nishimura, Harsha Prasada Lashkari, Sadashiva Rao, Stephanie Richards, Ji Eun Han, Ottavia M. Delmonte, Camille Kergaravat, Janet G. Markle, Masato Ogishi, Jing Han, Jessica Peel, Joseph Vellutini, Yi Feng, Camille Soudée, Mélanie Migaud, Boaz Palterer, Katherine J L Jackson, Shiho Nishimura, Sonoko Sakata, Keishiro Kinoshita, Ayako Yamamoto, Hiroshi Moritake, Mohammed Alzahrani, Francisco Vallejos, Theresa L Cole, Joanne M Smart, Sharon Choo, Zahra Chavoshzadeh, Shahnaz Armin, Antoine Toubert, Peng Zhang, Jeremie Rosain, Luigi D. Notarangelo, Qiang Pan-Hammarstrom, Stuart G. Tangye, Jean-Laurent Casanova, Cindy S. Ma, Anne Puel, Jacinta Bustamante, Satoshi Okada, Stephanie Boisson-Dupuis, Rui Yang

## Abstract

We previously reported inherited RORγT deficiency in seven patients from three ancestries (Chilean, Palestinian, Saudi Arabian) with mycobacterial disease and chronic mucocutaneous candidiasis (CMC). We report here five additional patients from different ancestries (Afghan, Indian, Iranian, Japanese, Sri Lankan), each homozygous for a new loss-of-function *RORC* variant. All but one patient — the exception receiving early prophylaxis — developed mycobacterial disease due to a near-complete depletion of innate-like adaptive T cells, including MAIT and iNKT cells, low counts of adaptive T_H_1* and CD8^+^ T cells, and impaired *Mycobacterium*-induced IFN-γ production by the remaining cells of these subsets, NK cells, conventional CD4^+^ T, Vδ1, and Vδ2 γδT cells. Most patients also displayed CMC due to their low counts of T_H_17 and T_H_1* cells. One patient died from disseminated Bacille Calmette-Guérin (BCG) vaccine infection, but, unexpectedly, all the other patients are still alive and clinically stable. RORγT is essential for protective immunity against mycobacteria and *Candida* in humans.

## Introduction

The clinical outcome of any infection varies considerably between individuals, ranging from asymptomatic infection in most to fatal disease in a few (Casanova, 2025; Casanova, 2015a; Casanova, 2015b; Casanova and Abel, 2021; Casanova and Abel, 2022). Tuberculosis (TB), caused by *Mycobacterium tuberculosis* (*Mtb*), is one of the deadliest diseases, accounting for at least one billion deaths in the past two millennia (Furin et al., 2019). The interindividual variability observed upon infection with *Mtb* is an enigma of biological and medical importance. Twin studies in the 1930s revealed a strong human genetic basis for TB susceptibility (Comstock, 1978; Kallmann and Reisner, 1943; Puffer, 1945). Studies of Mendelian susceptibility to mycobacterial disease (MSMD), an inherited, selective predisposition to clinical disease caused by weakly virulent mycobacteria, such as *Mycobacterium bovis* Bacille Calmette-Guérin (BCG) vaccines and environmental mycobacteria (EM), have paved the way for the molecular study of TB susceptibility (Boisson-Dupuis, 2020; Bustamante, 2020; Casanova and Abel, 2022). MSMD occurs in approximately 1 in 50,000 individuals. It can be categorized as “isolated” or “syndromic”. Isolated MSMD refers to a narrow phenotype characterized by selective vulnerability to weakly virulent mycobacteria and, occasionally, other intramacrophagic microorganisms such as *Salmonella*. By contrast, syndromic MSMD includes additional clinical infectious and non-infectious phenotypes. Both isolated and syndromic MSMD display a high level of genetic and allelic heterogeneity, with 47 genetic etiologies resulting from germline mutations in 22 genes (Bohlen et al., 2023; Casanova and Abel, 2022; Philippot et al., 2023; Rosain et al., 2023). Remarkably, MSMD displays biological homogeneity, as all the genes implicated in this condition encode proteins involved in interferon gamma (IFN-γ)-dependent immunity, with the possible exception of *ZNFX1* (Bohlen et al., 2023; Bustamante, 2020; Casanova and Abel, 2020; Casanova and Abel, 2022; Ogishi et al., 2023b). Disorders of *IFNG*, *IL12B*, *IL12RB1*, *IL12RB2*, *IL23R*, *TYK2*, *ISG15*, *RORC*, *MCTS1* and *TBX21* impair IFN-γ production, whereas disorders of *IFNGR1*, *IFNGR2*, *STAT1*, *JAK1*, *CYBB*, and *USP18* impair cellular responses to IFN-γ. Disorders of *IKBKG*, *SPPL2A*, *IRF1* and *IRF8* impair both the production of and responses to IFN-γ (Bohlen et al., 2023; Casanova and Abel, 2022; Philippot et al., 2023; Rosain et al., 2023). Inherited CCR2 deficiency reduces the recruitment of IFN-γ-responsive monocytes to the lungs and infected tissues, thereby underlying a syndromic form of MSMD with a complex pulmonary disease (Neehus et al., 2024).

Inherited deficiencies of two human transcription factors, T-bet and retinoic acid-related orphan receptor gamma T (RORγT), underlie syndromic MSMD (Okada et al., 2015; Yang et al., 2022; Yang et al., 2020; Yang et al., 2021). This finding was unexpected, as the corresponding mouse orthologs control the lineage commitment of two different T helper subsets, T_H_1 and T_H_17, respectively (Ivanov et al., 2006; Szabo et al., 2000). Autosomal recessive (AR) T-bet deficiency due to homozygosity for a *TBX21* variant underlies disseminated BCG infection (BCG-osis) and persistent airway hyperresponsiveness (Yang et al., 2020; Yang et al., 2021), whereas inherited RORγ/RORγT deficiency due to biallelic loss-of-function (LOF) *RORC* variants was found to underlie susceptibility to mycobacteria in seven patients from three ancestries (Chilean, Palestinian, and Saudi Arabian) (Okada et al., 2015). Specifically, inherited RORγT deficiency caused BCG-osis in six patients and disseminated TB in one patient, and chronic mucocutaneous candidiasis (CMC) in six of the seven patients (Okada et al., 2015). AR T-bet deficiency underlies mycobacterial susceptibility by severely impairing the development of innate (NK) and innate-like adaptive lymphocytes, including mucosa-associated invariant T (MAIT), invariant natural killer T (iNKT), and Vδ2^+^ γδ T cells, and the capacity of the remaining cells to produce IFN-γ. CCR6^−^ classical T_H_1 cells, which do not respond to mycobacterial infections in humans (Acosta-Rodriguez et al., 2007), are depleted in AR T-bet deficiency (Yang et al., 2020). By contrast, *Mycobacterium*-specific CD8^+^ and CD4^+^ T cells, including T_H_1* cells (Okada et al., 2015) — a subset of memory CD4^+^ T_H_1 cells to which almost all *Mycobacterium*-specific CD4^+^ T cells belong (Acosta-Rodriguez et al., 2007) — are capable of producing IFN-γ normally in inherited T-bet deficiency (Yang et al., 2020). However, the mechanism by which RORγ/RORγT governs antimycobacterial immunity is not fully understood (Okada et al., 2015). Patients with inherited RORγ/RORγT deficiency lack MAIT and iNKT cells and have impaired IFN-γ production by γδ T cells and T_H_1* cells (Okada et al., 2015). In addition, inherited RORγ/RORγT deficiency causes CMC of variable severity by impairing the production of IL-17 production, primarily by CD4^+^ αβ T cells, whereas, in AR T-bet deficiency, IL-17 production remains intact across lymphocyte subset (Okada et al., 2015; Yang et al., 2020). There have been no additional studies of inherited RORγ/RORγT deficiency or reports of new cases since our 2015 report (Okada et al., 2015). Here, we characterized human RORγT-dependent immunity in more detail by 1) reporting new patients with previously unknown or validated pathogenic variants of *RORC*; 2) updating the clinical phenotypes of previously reported patients; and 3) comprehensively dissecting the molecular and cellular mechanisms by which RORγT controls protective immunity.

## Results

### Updated clinical phenotypes of previously reported patients with RORγ/RORγT deficiency

We followed the seven previously reported patients (P1 – P7) with inherited RORγ/RORγT deficiency (Okada et al., 2015). Patients P1 – P3 (Kindred A) were born to consanguineous parents of Palestinian origin (**Fig. 1A** and **Table 1**). P1 died from BCG-osis before the age of 10 years. Both P2 and P3 developed BCG-osis; P1 and P2 also experienced recurrent oral candidiasis. Unfortunately, we were unable to maintain long-term follow-up for this kindred. It is therefore unknown whether P3 has developed CMC since our 2015 report (Okada et al., 2015). Patient P4 (Kindred B), a girl born to consanguineous Chilean parents, had a history of BCG-osis, recurrent oral thrush, vulvovaginal candidiasis, intertrigo, and onychomycosis. She has remained clinically stable on a regimen of isoniazid, trimethoprim-sulfamethoxazole (TMP-SMX) prophylaxis, and immunoglobulin replacement therapy (IgRT), initiated at presentation on the basis of a presumed diagnosis of severe combined immunodeficiency (SCID) or combined immunodeficiency (CID). This treatment has never been stopped (**Fig. 1A** and **Table 1**). Patients P5 – P7 (Kindred C) were born to consanguineous Saudi Arabian parents. P6 and P7 developed BCG-osis, whereas P5 experienced extrapulmonary TB. Since our initial report (Okada et al., 2015), all three have remained free from mycobacterial infections. Recurrent oral thrush in P5 – P7 has resolved, and onychomycosis in P5 and P6 has cleared. Recurrent intertrigo has resolved in P7 but continues to affect P6. P7 developed verrucae on both feet and common warts on the hands, which resolved following cryotherapy and topical salicylic acid treatment. P7 was also diagnosed with acute T-lymphoblastic leukemia (T-ALL), treated by chemotherapy, and is currently in remission (**Fig. 1A and Table 1**). All three patients (P5 – P7) have chronic hepatitis B; P6 and P7 have been receiving tenofovir. Both P5 and P6 have experienced recurrent dacryocystitis and dacryostenosis, requiring lacrimal duct stenting. P5, P6 and P7 are not currently on antimicrobial prophylaxis (**Fig. 1A** and **Table 1**). Thus, four of the previously reported patients living with inherited RORγ/RORγT deficiency that we have followed remain clinically stable, with or without appropriate antimycobacterial and antifungal prophylaxis, and they present only mild manifestations of CMC.

**Figure 1.**
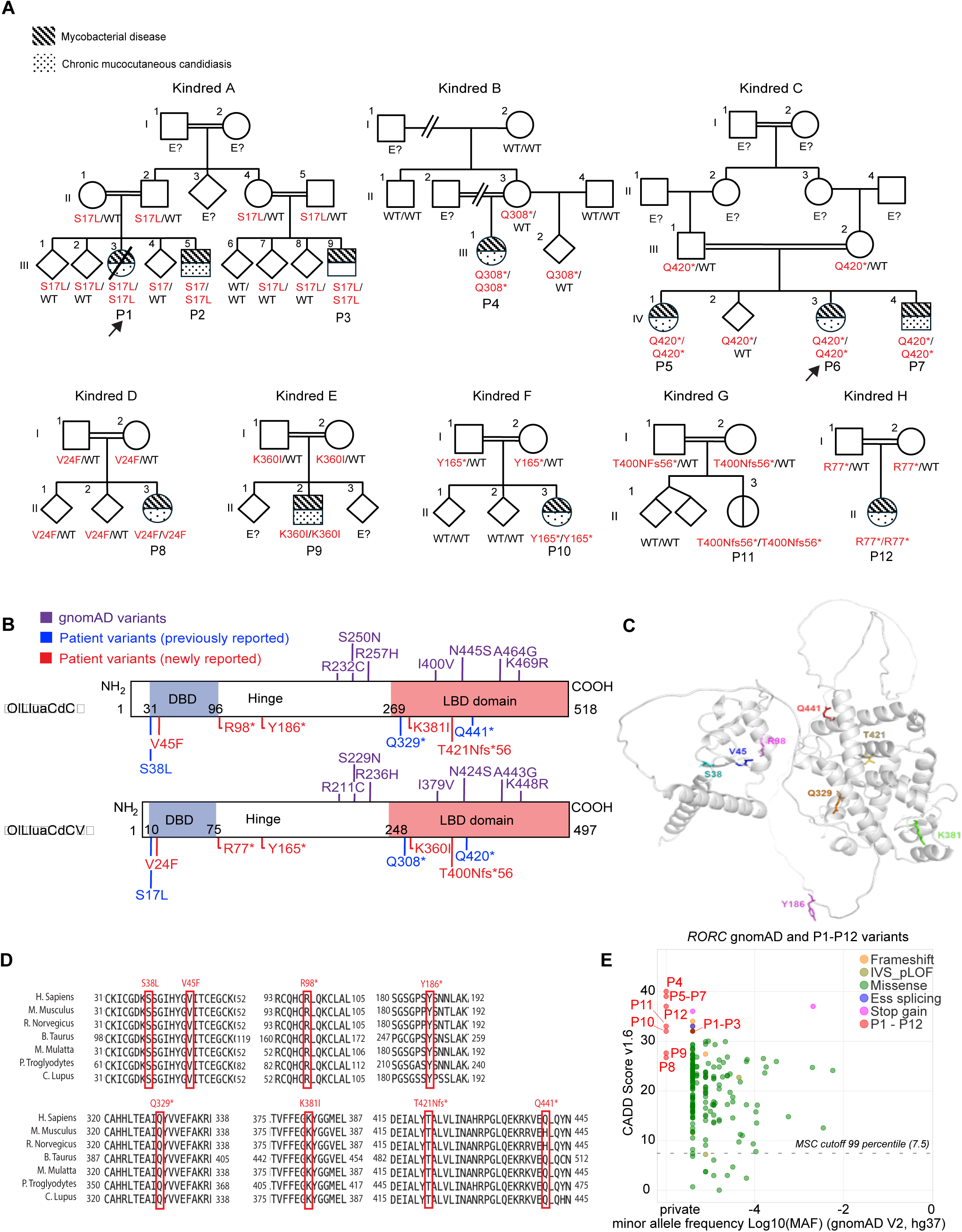
Twelve patients homozygous for *RORC* variants. (**A**) Pedigrees showing familial segregation of *RORC* variants. Shaded symbols indicate individuals with mycobacterial disease, and dotted symbols indicate individuals with chronic mucocutaneous candidiasis. (**B**) Schematic representation of RORγ and RORγT protein structures and their domains, encoded by *RORC* isoforms 1 and 2, respectively, together with the indicated variants. (**C**) Three-dimensional crystal structure of RORγ showing the locations of patient variants (V45F, R98*, Y186*, K381I, T421N*fs56). (**D**) Conservation of the amino-acid residues modified by the mutations across multiple species. (**E**) Graph of CADD v1.6 scores against minor allele frequencies (MAFs) showing the variants found in P1-P12, along with homozygous and heterozygous variants from gnomAD v2 (GRCh37).

**Table 1:**
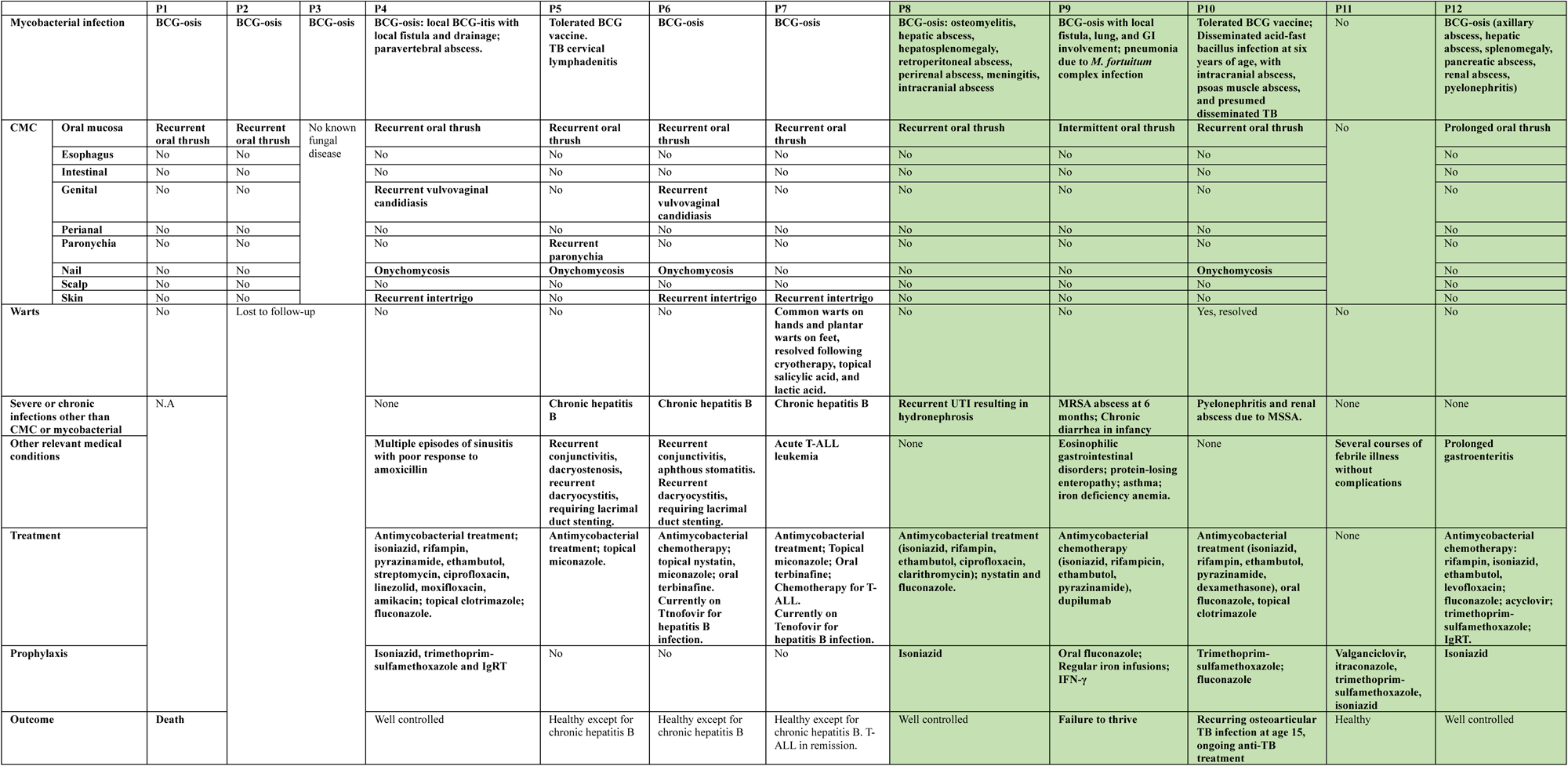
Clinical summary of previously published and new patients with autosomal recessive RORγ/RORγT deficiency. CMC: chronic mucocutaneous candidiasis. BCG-osis: disseminated Bacillus Calmette-Guérin (BCG) infection. IgRT: Ig replacement therapy. MRSA: methicillin-resistant *S. aureus* MSSA: methicillin-sensitive *S. aureus.* TB: tuberculosis. N.A: not applicable. T-ALL: T-cell acute lymphoblastic leukemia.

### Identification of five new unrelated patients with RORγ/RORγT deficiency

We studied five newly identified patients from unrelated consanguineous families residing in five different countries. Patient P8 (**Fig. 1A**, **Kindred D**, **Method case report**, and **Table 1**), a girl born to consanguineous parents of Afghan origin living in Iran, developed BCG-osis with chronic osteomyelitis, and intra-abdominal and intracranial abscesses following BCG vaccination. She also presented with recurrent oral candidiasis and T-cell lymphopenia (**Table 2**). Her condition was well managed with an antimycobacterial regimen and isoniazid prophylaxis (**Table 1**). Patient P9 (**Fig. 1A, Kindred E, Method case report,** and **Table 1**), a boy born to consanguineous Sri Lankan parents living in Australia, developed BCG-osis involving the gastrointestinal (GI) and respiratory tracts following BCG vaccination. He later presented with refractory pneumonia due to *M. fortuitum* complex, peripheral hypereosinophilia, eosinophilic gastrointestinal disorder (EGID) with protein-losing enteropathy (PLE), iron deficiency anemia (IDA), and asthma. EGID, PLE, and IDA improved following treatment with anti-IL-4Rα antibody (dupilumab). P9 also displayed mild intermittent mucocutaneous *Candida* infection, for which he receives antifungal prophylaxis. His mycobacterial disease has been controlled by regular recombinant IFN-γ infusions (**Table 1**). Patient P10 (**Fig. 1A, Kindred F, Method case report** and **Table 1**), a girl born to consanguineous Indian parents living in a region in which TB is endemic, developed intracranial and intra-abdominal abscesses due to disseminated infection with an acid-fast bacillus presumed to be *Mtb*. She displayed no adverse reaction to BCG vaccination at birth. She experienced refractory oral candidiasis, progressive onychomycosis, and widespread cutaneous warts, all of which were self-resolving. Her condition remained well controlled on fluconazole and TMP-SMX prophylaxis until adolescence, when she developed left-knee osteoarticular tuberculosis with *Mtb* arthritis, osteomyelitis, and intraosseous abscesses. At the time study she is still on an anti-TB regimen. Patient P11 (**Fig. 1A, Kindred G, Method case report,** and **Table 1**), a girl born to consanguineous Japanese parents, was identified on newborn screening as having undetectable levels of T-cell receptor excision circles (TRECs). She was subsequently diagnosed with T-cell lymphopenia (**Table 2**) and was not vaccinated with BCG. Apart from several febrile illnesses without complications, she has remained clinically asymptomatic whilst on valganciclovir, itraconazole, TMP-SMX, and isoniazid prophylaxis. Finally, patient P12 (**Fig. 1A, Kindred H, Method case report,** and **Table 1**), a girl born to consanguineous parents of Iranian origin, developed BCG-osis following vaccination, with liver, renal, and pancreatic abscesses and pyelonephritis. Immunological evaluation revealed T-cell lymphopenia and an absence of TREC detection (**Table 2**). P12 also had recurrent oral candidiasis and chronic diarrhea and has remained clinically stable on isoniazid prophylaxis.

**Table 2:**
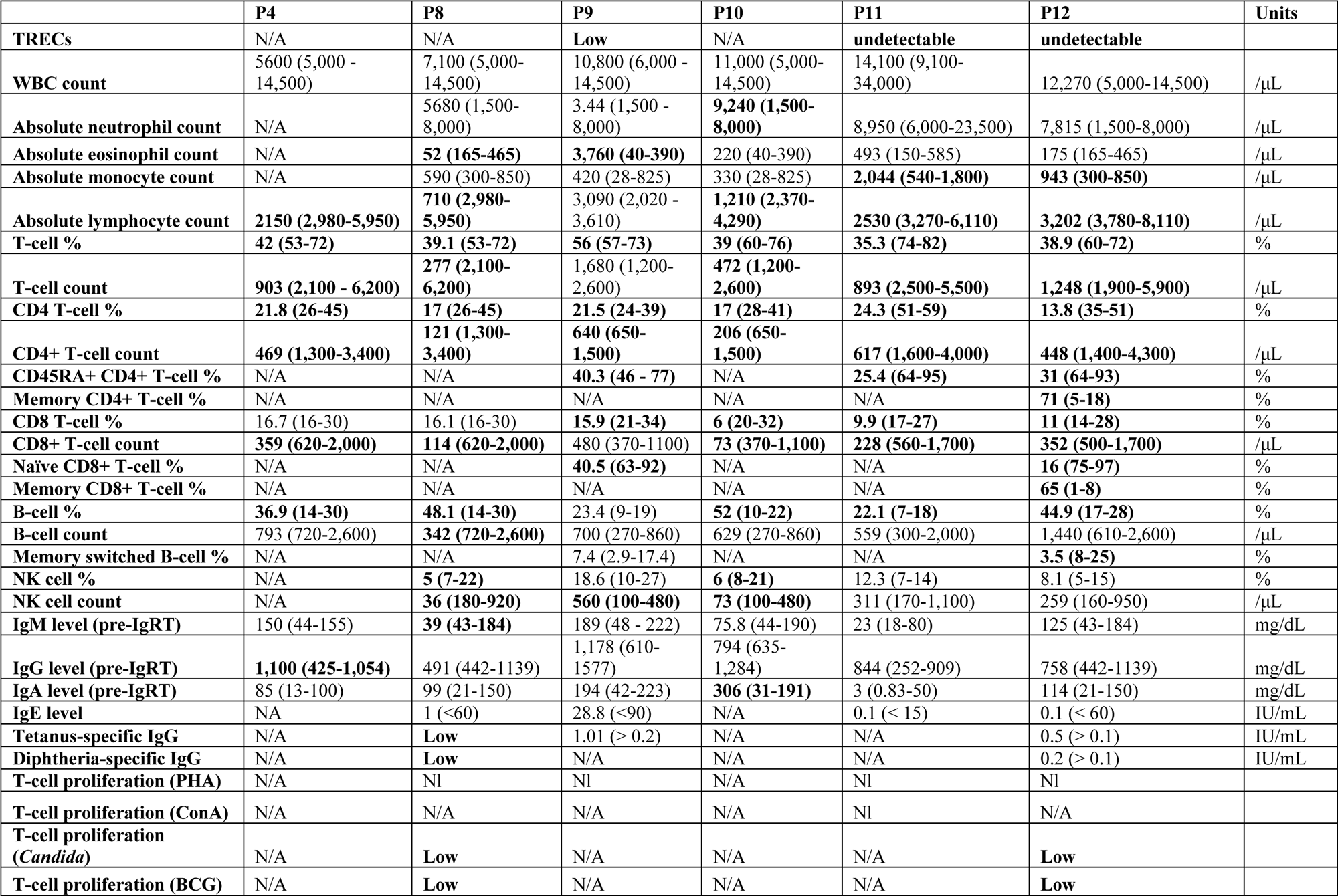
Clinical immunophenotypes of previously published and new patients with autosomal recessive RORγ/RORγT deficiency. TREC: T-cell receptor excision circles (reference range lab-specific). T cell %: CD3^+^ T % among lymphocytes. CD4 T-cell %: CD4^+^ T % among lymphocytes. CD45RA^+^ CD4^+^ T-cell %: CD45RA^+^ CD4^+^ T-cell % among CD4^+^ T cells. Memory CD4^+^ T-cell %: CD45RO^+^ % among CD4^+^ T cells. CD8 T-cell %: CD8^+^ T % among lymphocytes. Naïve CD8^+^ T-cell %: CD45RA^+^ % among CD8^+^ T cells. Memory CD8^+^ T-cell %: CD45RO^+^ among CD8^+^ T cells. B-cell %: CD19^+^ % among lymphocytes. Memory switched B-cell %: CD27^+^IgD^−^ B cells % among B cells. NK cell %: CD16^+^/CD56^+^ % among lymphocytes. PHA: phytohemagglutinin A. ConA: concanavalin A.

### Identification of five new *RORC* genotypes

We performed whole-exome sequencing (WES) on these five new patients. As all five patients were born to consanguineous parents and their first-degree relatives were reportedly healthy, we tested the hypothesis of an AR disorder due to homozygous variants. We prioritized homozygous variants that either had a minor allele frequency (MAF) below 0.003 in gnomAD v3 or were private (Chen et al., 2022; Karczewski et al., 2020), satisfying the following conditions: 1) non-synonymous exonic variants or variants affecting essential splicing sites; 2) variants with a combined annotation depletion-dependent (CADD) score above the 95th percentile of the mutation significance cutoff (MSC) (Itan et al., 2016; Kircher et al., 2014); 3) variants of genes with a gene damage index (GDI) below the cutoff of 13.36, which is tailored to AR inheritance (Itan et al., 2015); and 4) variants excluded from Blacklist, an in-house variant list corresponding to signals that are probably false positives (Maffucci et al., 2019). Our pipeline identified 19, 17, 15, 4, and 3 single-nucleotide variants (SNVs) satisfying these criteria in P8 – P12, respectively (**Fig. S1A**). *RORC* was the only gene for which rare variants were identified in all five patients. None of these five *RORC* variants had ever been reported. The consensus negative selection score (CoNeS) for *RORC* is −1.48, the 11^th^ lowest value of any gene causing AR IEI, suggesting that *RORC* is under strong negative selection (Rapaport et al., 2021). In humans, *RORC* generates two isoforms —isoform 1 or RORγ, and isoform 2 or RORγT — via the initiation of transcription from alternative start sites (Eberl and Littman, 2003; He et al., 1998; Medvedev et al., 1997; Ruan et al., 2011; Villey et al., 1999). P8 carried a homozygous missense variant within the DNA-binding domain (DBD) (c.133G>T, resulting in p.V45F for RORγ and p.V24F for RORγT), the AlphaMissense score of which was 0.9581, predicting pathogenicity (Cheng et al., 2023) (**Fig. 1A and B**). P9 was homozygous for a missense variant within the ligand-binding domain (LBD) (c.1142A>T, resulting in p.K381I for RORγ and p.K360I for RORγT) (**Fig. 1A and B**). The AlphaMissense score for K381I was also high, at 0.9025 (Cheng et al., 2023). P10 had a homozygous nonsense variant (c.558T>G) within the hinge domain (p.Y186* for RORγ and p.Y165* for RORγT) (**Fig. 1A and B**). P11 was homozygous for an insertion variant (c.1261_1262insA) predicted to result in a frameshift mutation due to a premature stop codon (p.T421Nfs56* for RORγ and p.T400Nfs*56 for RORγT) (**Fig. 1A and B**). Finally, P12 was homozygous for a nonsense variant (c.292C>T, p.R98* for RORγ and p.R77* for RORγT) within the hinge domain (**Fig. 1A and B**). Sanger sequencing confirmed the genotypes of all five patients (**Fig. S1B**). All parents were heterozygous carriers, and the healthy siblings of P8, P10, and P11 were either heterozygous or homozygous WT (**Fig. 1A** and **Fig. S1B**). Together with the three disease-causing variants reported in our previous study (Okada et al., 2015), these eight *RORC* variants result in amino-acid changes affecting all three domains of RORγ/RORγT (**Fig. 1B and 1C**). All eight amino acids affected by the variants are conserved across multiple species (**Fig. 1D**). The V45F, K381I, Y186*, T421Nfs*56, and R98* variants had CADD scores of 26.7, 27.6, 32, 33, and 37, respectively, according to CADD version 1.6 (Kircher et al., 2014; Zhang et al., 2018), all well above the 95^th^ percentile of the MSC of 7.5. R98* was reported in one heterozygous individual of Middle Eastern descent and one of European descent in gnomAD v4.1. By contrast, V45F, K381I, Y186*, and T421Nfs56* were not found in gnomAD v2.1, v3, v4 (Chen et al., 2022; Karczewski et al., 2020), Bravo (Taliun et al., 2021), Middle East cohort (Scott et al., 2016), or our in-house database of more than 20,000 exomes (**Fig. 1E**) (Zhang et al., 2018). Together, these data strongly suggest that these private homozygous *RORC* variants carried by P8 – P12 are probably deleterious and underlie the susceptibility to mycobacterial and/or fungal infections observed in these patients.

### The five mutant RORγ/RORγT alleles are LOF

We investigated the functional impact of the newly identified *RORC* variants by overexpressing cDNAs encoding WT or mutant *RORC* (both isoforms - isoform 1 (RORγ) and isoform 2 (RORγT)). Overexpression in HEK 293T cells resulted in similar levels of *RORC* RNA for the WT and all mutant constructs (**Fig. S1C**). The WT RORγ and RORγT proteins were predominantly localized to the nucleus (**Fig. S1D** and **Fig. 2A**). The expression and localization patterns of previously described variants — Q329*/Q308*, Q441*/Q420*, and S38L/S17L — were consistent with prior findings (Okada et al., 2015). The missense variants S38L/S17L, V45F/V24F, and K381I/K360I generated lower levels of a protein of the expected molecular weight, suggesting a lower stability of these mutant forms (**Fig. S1D** and **Fig. 2A**). By contrast, the nonsense variants Q329*/Q308*, Q441*/Q420*, and Y186*/Y165* generated only very small amounts of truncated RORγ/RORγT proteins. No protein was detected for the R98*/R77* variant (**Fig. S1D** and **Fig. 2A**). The T421Nfs*56/T400Nfs*56 frameshift variant generated low levels of a truncated protein (**Fig. S1D** and **Fig. 2A**). All variants – except for R98*/R77*, for which no protein was produced – were predominantly retained in the nucleus. We then investigated the DNA-binding capacity of these mutants. WT RORγ and RORγT bound ROR response elements (RORE) efficiently, but both the previously reported S38L/S17L, Q329*/Q308* and Q441*/Q420* variants, and the newly identified V45F/V24F, K381I/K360I, T421Nfs*56/T400Nfs*56, and R98*/R77* variants failed to bind RORE (**Fig. 2B** and **Fig. S1E**). Interestingly, the truncated Y186*/Y165* mutant proteins from P10 had a lower molecular weight and displayed enhanced RORE binding (**Fig. 2B** and **Fig. S1E**). We investigated transcriptional activity further with an *in vitro* luciferase reporter system under the control of multimerized RORE (Ecoeur et al., 2019). The overexpression of WT RORγ/RORγT induced high levels of luciferase activity under the control of WT-RORE but not mutant-RORE (**Fig. 2C** and **Fig. S1F**). As expected (Okada et al., 2015), the overexpression of known LOF alleles — S38L/S17L, Q329X/Q308X, and Q441*/Q420* — did not induce a luciferase signal (**Fig. 2C** and **Fig. S1F**). Overexpression of the newly identified variants — V45F/V24F, K381I/K360I, Y186*/Y165*, T421Nfs*56/T400Nfs*56 and R98*/R77* *RORC —*abolished transcriptional activity (**Fig. 2C** and **Fig. S1F**). *IL17A* expression was robustly induced by overexpression of the WT *RORC* isoform 2 in Jurkat T cells, but not by any of the previously reported (S17L, Q308*, Q420*) or new (V24F, K360I, T400Nfs*56, and R77*) variants (**Fig. 2D**). The V45F/V24F, K381I/K460I, Y186*/Y165*, T421Nfs*56/ T400Nfs*56, and R98*/R77* *RORC* variants were therefore LOF when overexpressed.

**Figure 2.**
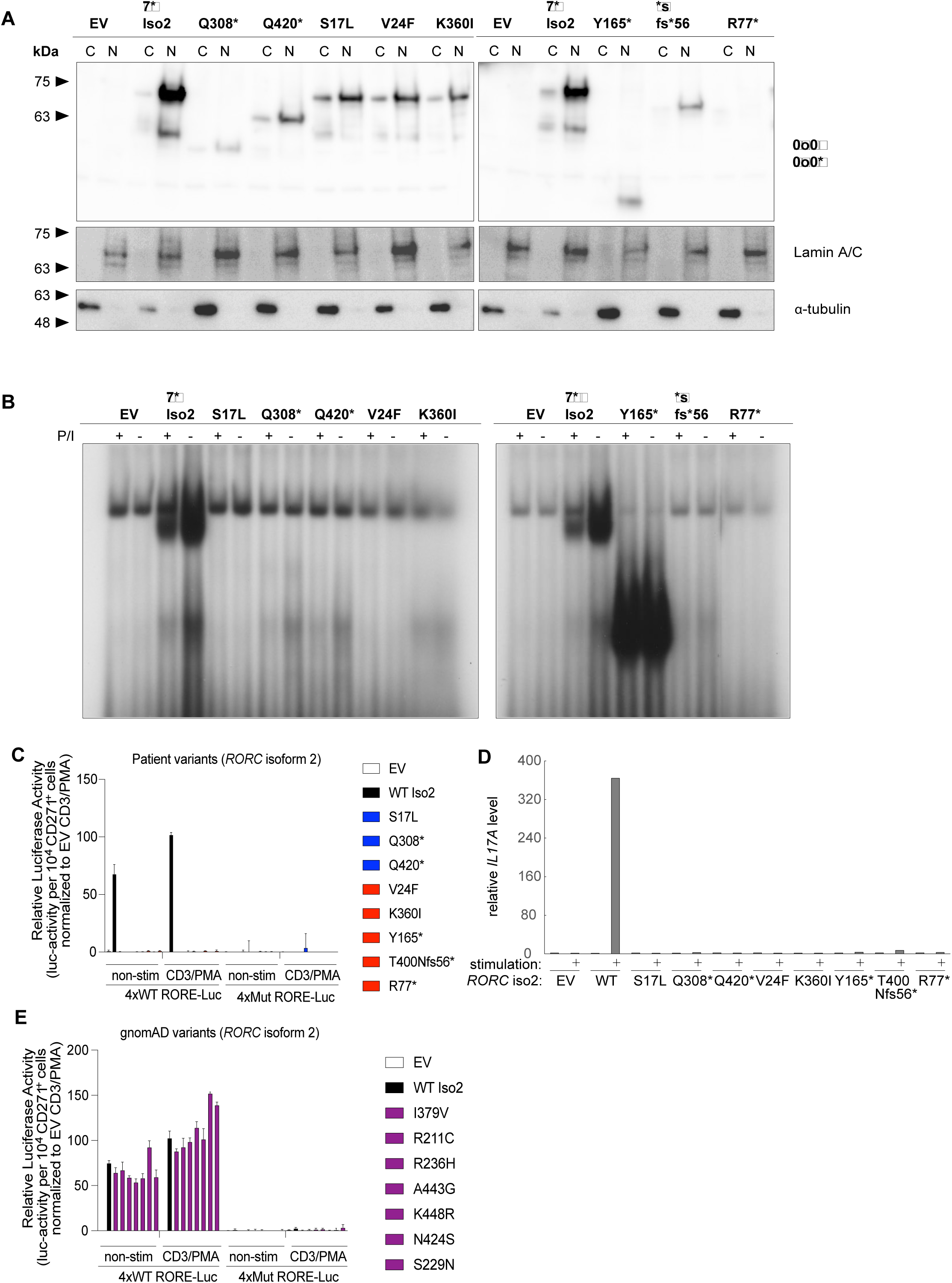
Functional characterization of mutant *RORC* isoform 2 variants under overexpression conditions. (**A**) Western-blot analysis of patient mutant *RORC* isoform 2 (RORγT) variants to assess protein levels and subcellular distribution in HEK293T cells. C, cytoplasmic fraction; N, nuclear fraction. (**B**) Electromobility shift assay (EMSA) with a ^32^P-labeled RORE-2 probe derived from the *IL17A* promoter, incubated with nuclear lysates of HEK239T cells transfected with the indicated vectors encoding RORγT, with or without PMA/ionomycin stimulation (PMA/Iono). (**C**) Transcriptional activity of patient RORγT variants in an *in vitro* luciferase reporter system driven by WT or mutant multimerized RORE. (**D**) *IL17A* mRNA levels relative to *GAPDH* mRNA levels in Jurkat T cells overexpressing WT or mutant *RORC* isoform 2, as indicated. (**E**) Transcriptional activity of common *RORC* variants from gnomAD v2.1 and 3.1, assessed as in (C). All experimental data were verified in at least 2 independent experiments.

### Population genetics supports the pathogenicity of *RORC* mutations

In gnomAD v2.1, v3, and v4.1 (Chen et al., 2022; Karczewski et al., 2020), eight variants — I400V, R232C, R257H, A464G, K469R, N445S, S250N and R10* — of isoform 1 RORγ, with MAF of 4.7 ×10^−5^, 1.4×10^−5^, 3.3×10^−5^, 1.3×10^−3^, 2.7×10^−4^, 4.5×10^−3^, 2.7×10^−3^, respectively, were present in the homozygous state in the general population (**Fig. 1B and 1E**). With the exception of A464G, all these variants had CADD scores above the 95^th^ percentile of the MSC (Itan et al., 2016). R10* is predicted to result in a truncated RORγ, but is not expected to affect RORγT, the transcription site of which lies more than 5 kb downstream from this variant. We evaluated the transcriptional activity of all variants except R10*, which does not affect the transcription or translation of RORγT. Specifically, we tested I400V/I379V, R232C/R211C, R257H/R236H, A464G/A443G, K469R/K448R, N445S/N424S, and S250N/S229N. Overexpression of the WT and these variant *RORC* alleles resulted in similar levels of protein, for both isoforms (**Fig. S1G and S1H**). The WT and mutant alleles also yielded similar and sufficient levels of luciferase activity driven by WT-RORE, but not mutant-RORE (**Fig. 2E** and **Fig. S1E**). Thus, all *RORC* variants encoding RORγT present in the homozygous state in the general population appear to be functionally neutral. In total, 25 predicted LOF (pLOF) *RORC* variants were found in gnomAD v4.1 (c.40+1G>A, c.14del, c.1535C>G, c.292C>T, c.41-2A>G, c.41-1G>A, c.70+2T>A, c.64del, c.211C>T, c.230_231insGTTT, c.244C>T, c.298+1G>A, c.273C>A, c.267del, c.1395+2T>C, c.410del, c.811+2_811+12del, c.1231del, c.997del, c.1432C>T, c.1530_1531del, c.598dup, c.1005_1006del, c.1126_1129del, c.646_647del), all exclusively in the heterozygous state (Chen et al., 2022; Karczewski et al., 2020). The cumulative MAF of these pLOF variants is approximately 2×10^−5^, corresponding to an estimated biallelic pLOF frequency of approximately 4×10^−10^, substantially lower than the known prevalence of MSMD and CMC. Thus, considering the genetic data for P8 – P12 and for previously published patients with AR RORγ/RORγT deficiency (P1 – P7) (Okada et al., 2015) and population genetics data, biallelic LOF *RORC* variants are exceedingly rare in the general population. The evidence provided by population genetics does not, therefore, contradict our hypothesis that the infectious and immunological phenotypes observed in P8 – P12 result from *RORC* deficiency.

### Homozygosity for LOF *RORC* variants impairs T-cell receptor (TCR)α rearrangement

RORγT is highly expressed in CD4^+^CD8^+^ (double positive, DP) thymocytes in mice and humans (Naik et al., 2024; Villey et al., 1999). RORγT deficiency restricts the T-cell repertoire by limiting TCRα gene usage in both species (Guo et al., 2016; Okada et al., 2015). In mice, the loss of RORγT function shortens the lifespan of DP thymocytes, thereby reducing the time window available for TCRα rearrangement (Guo et al., 2016; Okada et al., 2015). Furthermore, RORγT binds to a DP-specific component of the RAG antisilencer (DPASE), promoting the expression of *RAG* genes (Naik et al., 2024), and the activity of the T early α (TEA) promoter and a *Tcra* enhancer, increasing the accessibility of the *Tcra* locus (Naik et al., 2024; Villey et al., 1999). High-throughput sequencing of the *TRAV/TRAJ* loci in P8 – P12 revealed lower levels of usage for both the TCR-Vα and -Jα gene segments than were observed in the WT siblings of P11 (**Fig. 3A**). Specifically, the usage of distal *TRAV* segments, particularly *TRAV01-01* to *TRAV08-02*, was significantly lower in P8 – P12 than in healthy adult or age-matched controls (**Fig. 3A**). Similarly, distal TCR-Jα segments, particularly *TRAJ01* to *TRAJ34*, were underused in P8 – P12 (**Fig. 3A**). By contrast, an enrichment in proximally located *TRAV* and *TRAJ* segments — *TRAV38* to *TRAV41* and *TRAJ54* to *TRAJ61* in particular — was observed in P8 – P12 (**Fig. 3A**). This defect of distal TCR-Vα and -Jα rearrangement was not observed in any of the seven heterozygous parents carrying LOF *RORC* alleles or in healthy controls (**Fig. S2A**), suggesting a strictly recessive mode of inheritance for this phenotype. Two age-matched siblings of P11 had broad and diverse TCRα clonotypes (**Fig. 3B**). P11 and P12 had a clonotype diversity similar to that of controls (**Fig. 3B**). By contrast, P8, P9, and P10 had several dominant TCRα clonotypes and a low level of TCRα complementarity-determining region 3 (CDR3) diversity (**Fig. 3B**). This lower diversity in P8, P9, and P10 was further supported by a lower Shannon entropy, which measures overall repertoire diversity, and lower D50 values, corresponding to the proportion of unique clonotypes required to account for 50% of all TCRα reads, indicating a skewed TCRα repertoire (**Fig. 3C** and **Fig. 3D**). By contrast, P11 and P12 had entropy levels similar to those in controls, suggesting variable degrees of clonal diversity in patients with inherited RORγ/RORγT deficiency. In particular, P11 is the only patient never to have experienced severe or chronic infection; there is probably, therefore, little confounding of her TCRα profile. Remarkably, the clonotype diversity, Shannon entropy, and D50 of P11 were similar to those of both adult and age-matched sibling controls (**Fig. 3B–D**). Similar defects of TCRα rearrangement were also reported in P1 – P7 (Okada et al., 2015). Thus, RORγT is essential for optimal TCRα recombination, and for full usage of the distal *TRAV* and *TRAJ* segments in particular. However, RORγT does not appear to be required for the maintenance of CDR3 diversity.

**Figure 3.**
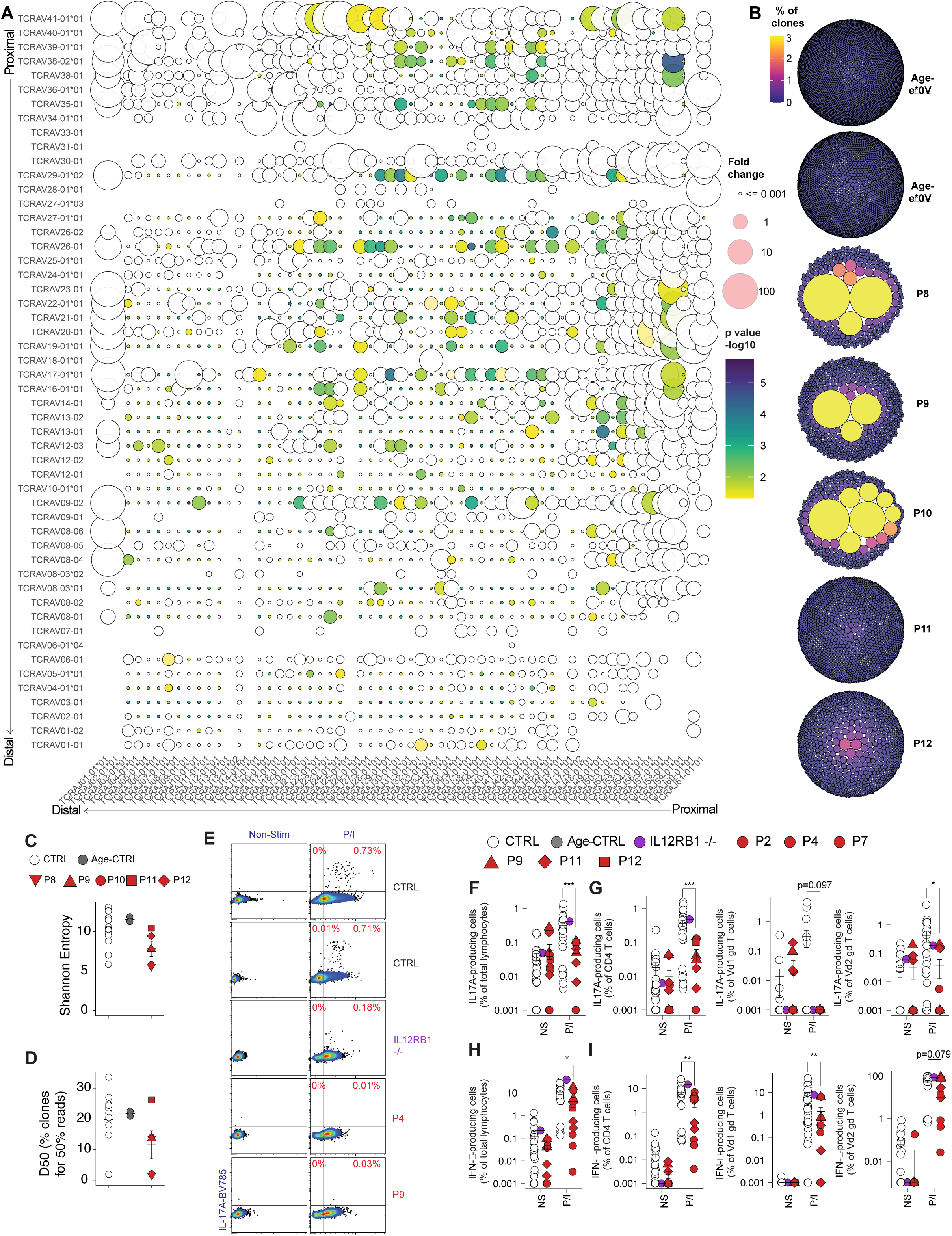
The endogenous RORγ/RORγT of patients are loss-of-function, as demonstrated by the impairment of TCRα rearrangement and abolition of IL-17A production. (**A**) Bubble plot of high-throughput sequencing of the *TRAV/TRAJ* loci of healthy controls and P8 – P12. The fold-change difference in *TRAV* and *TRAJ* usage in P8 – P12 relative to controls is indicated by bubble size, and statistical significance (*p*-value) by color, with *TRAV* segments on the *y*-axis and *TRAJ* segments on the *x*-axis. (**B**) Clonotype diversity representation for two age-matched controls and P8 – P12, with each mini-circle denoting one unique clonotype based on TCRα sequences. (**C**) Shannon entropy index of TCRα repertoires in healthy controls and P8 – P12, reflecting overall diversity. (**D**) D50 values, proportion of unique clonotypes accounting for 50% of total TCRα reads, in healthy controls and P8 – P12. (**E**) Dot-plots showing expression of RORγT and IL-17A in controls and patients, with or without PMA/iono (P/I) stimulation. (**F** and **G**) Frequency of IL-17A-producing cells among total lymphocytes (F) and within indicated subsets (G) in controls or patients, with or without P/I stimulation. (**H** and **I**) Frequency of IFN-γ-producing cells among total lymphocytes (H) and within indicated subsets (I) in controls or patients, with or without P/I stimulation. All experimental data were verified in at least 2 independent experiments.

### The patients’ endogenous RORγT is LOF

RORγT governs the lineage commitment of T_H_17 cells, a memory CD4^+^ T-cell subset characterized by an ability to secrete cytokines of the IL-17 family in both mice and humans, and to mediate immunity to fungal infections (Ivanov et al., 2006; Okada et al., 2015). We evaluated T_H_17 function in patients with *RORC* mutations by assessing *ex vivo* cytokine production by CD4^+^ T cells from patients. We investigated the reaction of lymphocytes from control individuals (CTRLs), previously reported patients P2, P4, and P7 (Okada et al., 2015), and new patients P9, P11, and P12 to stimulation with phorbol 12-myristate 13-acetate (PMA) and ionomycin (P/I). A patient with biallelic LOF variants of *IL12RB1* was included as a control. Approximately 0.5-1% of CTRL lymphocytes produced IL-17A, and these IL-17A-producing lymphocytes consistently expressed higher levels of RORγT than the majority of IL-17A-negative lymphocytes (**Fig. 3E and Fig. 3F**). By contrast, RORγ/RORγT-deficient patients had a markedly lower frequency of IL-17A-producing lymphocytes (**Fig. 3E and Fig. 3F**) attributed principally to effects on CD4^+^ T cells, Vδ1^+^ γδ T cells, and Vδ2^+^ γδ T cells (**Fig. 3G**), but not CD8^+^ T, NK, or B cells (**Fig. S2B**). Interestingly, a statistically significant but subtle decrease in IFN-γ production by lymphocytes was also observed in patients with RORγ/RORγT deficiency relative to CTRLs (**Fig. 3H**). This decrease was driven by the lower levels of IFN-γ production by CD4^+^ T cells, Vδ1^+^ γδ T cells, Vδ2^+^ γδ T cells, and B cells (**Fig. 3I** and **Fig. S2C**), but not by CD8^+^ T or NK cells (**Fig. S2C**). These data, together with the impaired TCRα rearrangement in P8 – P12, demonstrate that the endogenous RORγT expressed in P8 – P12 is LOF, and that the resulting inability of CD4^+^ T, Vδ1^+^ γδ T, and Vδ2^+^ γδ T cells to produce IL-17A in these patients underlies their susceptibility to mucocutaneous *Candida* infection.

### Normal myeloid cell development in patients with RORγ/RORγT deficiency

We investigated the role of RORγT in the development of leukocyte lineages by first analyzing the complete blood counts of all patients for which such counts were available. The absolute numbers of white blood cells (WBC), neutrophils, and monocytes were within the age-matched reference ranges, or at least not meaningfully outside them (**Table 2**). Absolute eosinophil count was unremarkable in all patients except P9, who developed peripheral hypereosinophilia at 3,760 cells per microliter and EGID (**Table 2**). We then performed comprehensive 40-color immunophenotyping of peripheral blood mononuclear cells (PBMCs) from healthy controls (CTRLs), two patients with AR IL-12Rβ1 deficiency — another IEI that underlies both MSMD and, to a lesser extent, CMC (Ouederni et al., 2014) — previously reported patient P4 (Okada et al., 2015), and new patients P9, P10, P11, and P12, by spectral flow cytometry (Ogishi et al., 2022). Unbiased clustering analysis revealed 13 major immune cell subsets (Ogishi et al., 2021), including both myeloid subsets — monocytes and dendritic cells (DC) — and lymphoid populations — CD4^+^ αβ T, CD8^+^ αβ T, double-negative (DN) T, Vδ1^+^ γδ, Vδ2^+^ γδ, MAIT, iNKT, NK, innate lymphoid cells (ILC), and B cells (**Fig. S3A**). The frequencies of classical, intermediate, and nonclassical monocytes were similar in CTRLs, age-matched CTRLs (age-CTRLs), and RORγ/RORγT-deficient patients (**Fig. S3A**). Similarly, frequencies of myeloid DC, including both conventional DC (cDC)1 and cDC2, and plasmacytoid DC (pDC) were also mostly similar between patients and CTRLs (**Fig. S3B and S3C**). Thus, the development of all major myeloid lineages remains intact in individuals with inherited RORγ/RORγT deficiency.

### Normal development of NK and B cells but impaired development of innate lymphoid cells

We then investigated the lymphoid compartment in individuals with RORγ/RORγT deficiency. Absolute lymphocyte counts (ALCs) were consistently low in all patients except P9 (**Table 2**). We analyzed the distribution of lymphocyte subsets to determine whether this lymphopenia reflected specific defects of lymphoid subsets. Total counts of peripheral blood B and NK cells did not differ significantly between CTRLs and patients with RORγ/RORγT deficiency (**Table 2**). Frequencies of total NK cells, including CD16^+^CD56^dim^ and CD56^bright^ subsets, were not affected by RORγ/RORγT deficiency (**Fig. S3D**). Similarly, frequencies of total B cells and naïve, memory and plasma cell subsets were similar between CTRLs and patients with RORγ/RORγT deficiency (**Fig. S3E and S3F**). Consistent with these findings, baseline serum IgG levels were within the age-appropriate normal range in all patients (**Table 2**); tetanus- and diphtheria-specific IgG levels were protective in P9 and P12, but low in P8, who had not completed tetanus and diphtheria primary series by the time of blood collection **(Table 2**), and none of the patients experienced recurrent otosinopulmonary infections, a hallmark of primary antibody deficiency (**Table 1**) (Ballow, 2002). Thus, humoral immunity appears to be intact in inherited RORγt deficiency. RORγT is expressed by all innate lymphoid cell (ILC) lineages (Scoville et al., 2016) and is required for lineage commitment to IL-17-producing ILC3s and their designated ILC3 progenitors in humans (Lim et al., 2017). Consistent with a previous report (Lim et al., 2017), CD117^+^ ILC progenitor (ILCP) cells were less frequent in patients with RORγ/RORγT deficiency than controls, despite their similar frequencies of total ILCs (**Fig. 4B and 4C**). However, the frequency of ILC2s remained similar in age-matched CTRLs and patients (**Fig. S3G**). Despite the crucial role of RORγT in the differentiation of ILC3 and lymphoid tissue inducer (LTi) cells from ILCP in both humans and mice (Cherrier et al., 2012; Eberl et al., 2004; Lim et al., 2017; Sawa et al., 2010; Serafini et al., 2015), investigations of ILC3 and ILC1 were limited by the low abundance of these cells in peripheral blood. A new subset of major histocompatibility complex (MHC)-II^+^ LTi-like ILCs with antigen-presenting cell features was recently discovered in mice (Abramson et al., 2023). However, these cells were also too scarce for detection among PBMCs from either CTRL or RORγ/RORγT-deficient patients. In summary, the development of NK and B cells remained intact but the development of ILCP cells was impaired in patients with inherited RORγ/RORγT deficiency.

**Figure 4.**
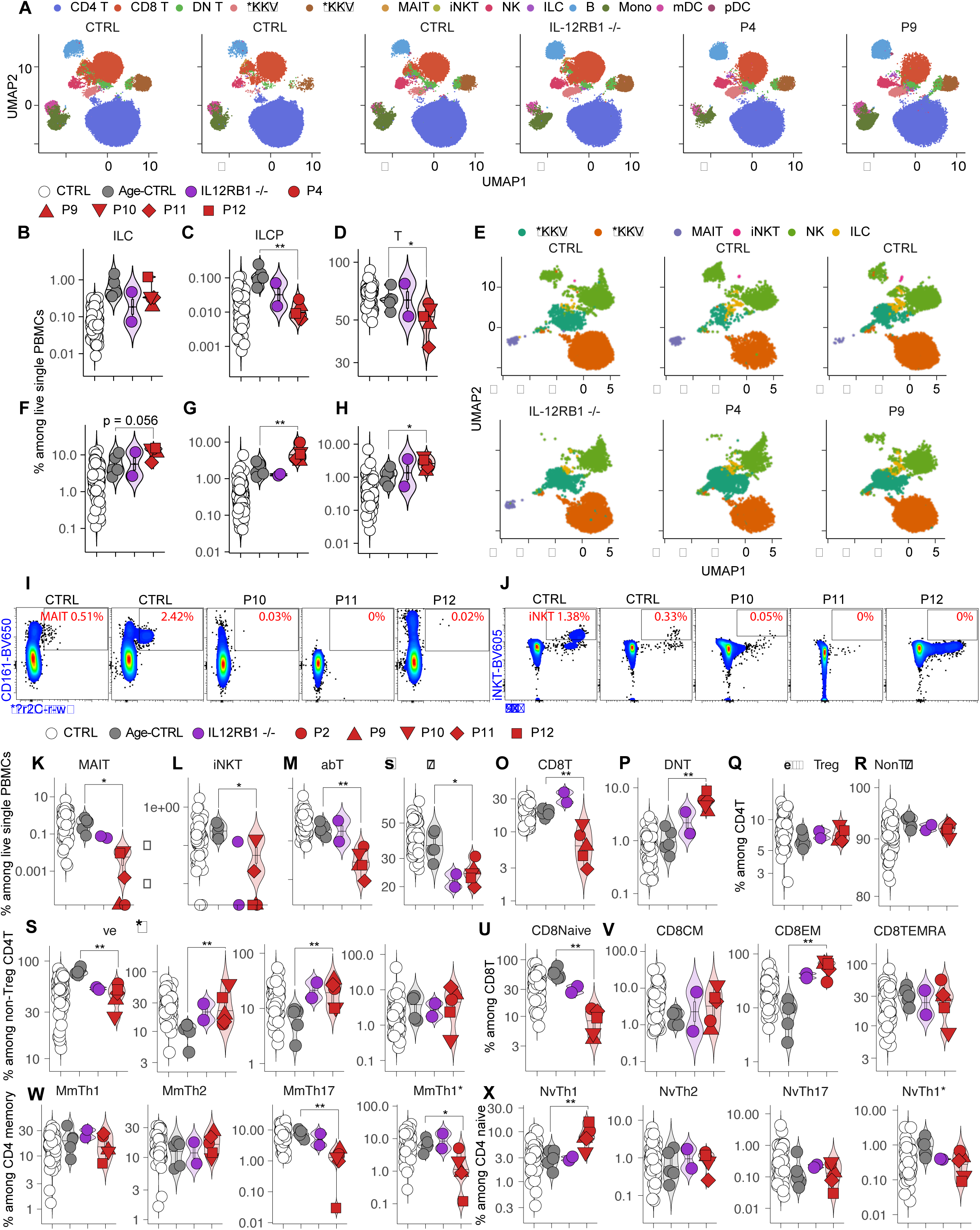
Deep immunophenotyping of patients with AR RORγ/RORγT deficiency. (**A**) Identification of major immune cell subsets in PBMCs from healthy controls and patients by unbiased clustering analysis. (**B - D**) Frequencies of innate lymphoid cells (ILCs) (B), ILC progenitors (ILCPs) (C), and T cells (D) among live PBMCs from controls, age-matched controls, and patients with AR IL-12Rβ1 or RORγ/RORγT deficiencies. (**E**) Visualization of major lymphoid subsets in healthy controls and patients by unbiased clustering. (**F - H**) Frequencies of γδ T (F), Vδ1^+^ γδ T (G), and Vδ1^−^Vδ2^−^ γδ T cells (H), among live PBMCs from controls, age-matched controls, and patients with AR IL-12Rβ1 or RORγ/RORγT deficiencies. (**I** and **J**) Dot-plots showing the presence of mucosal-associated invariant T (MAIT) (I) or invariant NKT cells (iNKT) cells (J). (**K – V**) Frequencies of MAIT (K), iNKT (L), αβ T (M), CD4^+^ T (N), CD8^+^ T (O), double-negative T (DNT) (P), CD4^+^ Treg (Q), naïve CD4^+^ T (R), memory CD4^+^ T (S), central memory (CM), effector memory (EM), T_EMRA_ CD4^+^ T (T), naïve CD8^+^ T (U), and CM, EM, T_EMRA_ CD8^+^ T (V) cells among live PBMCs from healthy controls or patients. (**W**) Frequencies of memory T_H_1, T_H_2, T_H_17, T_H_1* cells among memory CD4^+^ T cells. (**X**) Frequencies of naïve CD4^+^ T cells with T_H_1-, T_H_2-, T_H_17-, and T_H_1*-like phenotypes. All experimental data were verified in at least 2 independent experiments.

### Profound T-cell lymphopenia due to impaired thymopoiesis

Unlike NK and B cells, CD3^+^ T lymphocytes were significantly less frequent in all patients with inherited RORγ/RORγT deficiency than in controls, and absolute counts were lower in all patients except P9 (**Fig. 4D** and **Table 2**). This numerical deficiency of T cells accounted for the total lymphocyte counts below the lower limit of the age-matched reference range (**Table 2**). TRECs, circular δRec-ψJα DNA by-products of TCRα rearrangement, were undetectable in the peripheral blood of P11 and P12, and their levels were markedly low in P9, the only patients for whom data were available (**Table 2**). This finding is consistent with the impairment of TCRα rearrangement, as demonstrated above, in these patients. P11 is the only patient for whom T-cell counts from the neonatal period were available, with longitudinal T-cell data collected when the patient was in a healthy, infection-free state. At 12 days of age, her absolute T-cell count was low, at 893/µL (age-matched reference range: 2,500 – 5,500/µL) (**Table 2**) (Shearer et al., 2014). However, absolute T-cell count increased steadily over time and was only slightly below the 10^th^ percentile of the age-matched reference range by 19 months of age (**Fig. S3H**). RORγT function is essential for the temporal and spatial regulation of thymocyte development (Guo et al., 2016; Okada et al., 2015). RORγT promotes the survival of DP thymocytes by upregulating the expression of anti-apoptotic genes, and orchestrates both the intrathymic positioning and eventual emigration of maturing T cells (Guo et al., 2016). Collectively, these data suggest that *de novo* T-cell production is severely impaired at birth due to poor thymic output. However, as thymocytes continue to mature in the thymus and peripheral T cells undergo homeostatic proliferation, T-cell counts gradually rise over time. This pattern of delayed T-cell recovery is reminiscent of that observed in patients with other congenital thymic defects, such as haploinsufficiency of *FOXN1* or *FOXI3* (Bosticardo et al., 2019; Ghosh et al., 2022).

### Enrichment in blood γδ T cells and depletion of MAIT and iNKT cells

We further characterized the CD3^+^ T-cell compartment in inherited RORγ/RORγT deficiency by investigating both innate-like and conventional T-lymphocyte subsets. We first performed unbiased clustering to focus on innate-like adaptive T lymphocytes (**Fig. 4E**). A slight enrichment in total γδ T cells was observed in patients with RORγ/RORγT deficiency (**Fig. 4F**). It was attributed principally to an enrichment in Vδ1^+^ and Vδ1^−^Vδ2^−^ γδ T cells, but not Vδ2^+^ γδ T cells (**Fig. 4G, 4H and Fig. S3I**). This increase may result from skewed TCRα rearrangement favoring the TCRδ locus, which is nested within the *TRAV/TRAJ* loci (Manso et al., 2022). By contrast, MAIT and iNKT cells, the development of which is dependent on the correct arrangement of MHC-related protein-1 (MR1)-restricted Vα7.2-Jα33 and CD1d-restricted TCR-Vα24Jα18, respectively, was abolished in patients with RORγ/RORγT deficiency (**Fig. 4I–4L**, **Fig. S3J**). Notably, use of the distal *TRAV0102*, encoding TCR-Vα7.2, for MAIT cells, and *TRAJ18*, encoding TCR-Jα18, for iNKT cells was abolished and profoundly diminished, respectively, in all patients (**Fig. 3A**). The depletion of MAIT and iNKT cells confirms our previous findings reported at the time of the initial discovery of RORγ/RORγT deficiency (Okada et al., 2015). Similar findings were also obtained for an MSMD patient with AR T-bet deficiency (Yang et al., 2020), suggesting a non-redundant role of these two innate-like adaptive T-cell subsets in antimycobacterial immunity.

### Altered T helper cell differentiation in patients with RORγ/RORγT deficiency

We then investigated the development of conventional adaptive T lymphocytes in patients with inherited RORγ/RORγT deficiency. Absolute counts and frequencies of CD4^+^ and CD8^+^ T cells were low in all patients (**Table 2**, **Figure 4M–4O**). In P11, counts of both CD4^+^ and, to a lesser extent, CD8^+^ T cells tended to increase with age, with CD4^+^ T-cell counts approaching the 10^th^ percentile of the age-matched reference range by the age of 19 months (**Fig. S3K and S3L**). By contrast, the frequency of CD4^−^CD8^−^ double-negative (DN) T cells was markedly high in patients with inherited RORγ/RORγT deficiency (**Fig. 4P**), whereas the frequencies of regulatory T cells (T_REG_) and non-T_REG_ CD4^+^ T cells remained unaffected (**Fig. 4Q** and **4R**). Naive (CD45RA^+^CCR7^+^) CD4^+^ T-cell counts were low (**Fig. 4S**), whereas the central memory (CM) and effector memory (EM) CD4^+^ T-cell compartments were expanded in patients with RORγ/RORγT deficiency (**Fig. 4T**). The naïve CD4^+^ T-cell pool also expanded with age in P11 (**Fig. S3M**), suggesting a recovery of thymic output over time. The frequency of naïve CD8^+^ T cells was markedly low (**Fig. 4U**). By contrast, the EM CD8^+^ T-cell compartment was expanded (**Fig. 4V**). Despite the crucial role of T-bet in T_H_1 differentiation (Flynn and Chan, 2001; Szabo et al., 2000; Yang et al., 2020), a patient with inherited T-bet deficiency was found to retain IFN-γ production by CD4^+^ T cells during mycobacterial infections (Yang et al., 2020), suggesting that T_H_1 cells may, paradoxically, be redundant in antimycobacterial immunity. T_H_1 and T_H_2 frequencies were similar in CTRLs and RORγ/RORγT-deficient patients (**Fig. 4W**). By contrast, the frequency of memory T_H_17 cells, the development of which is governed by RORγT in mice (Ivanov et al., 2006), was significantly lower in patients with RORγ/RORγT deficiency than in controls (**Fig. 4W**), confirming the non-redundant role of RORγT in T_H_17 lineage commitment in humans. The frequency of memory T_H_1* cells was also lower in patients with RORγ/RORγT deficiency (**Fig. 4W**). CCR4^+^CCR6^+^CXCR3^−^ and CCR4^−^CCR6^+^CXCR3^+^ cells were preserved in the naïve CD4^+^ T-cell compartment, suggesting that the development of T_H_17 and T_H_1* cells may be dependent on antigen priming (**Fig. 4X**). However, there appeared to be an enrichment in naïve CD4^+^ T cells with a T_H_1 phenotype in patients with RORγ/RORγT deficiency (**Fig. 4X**). A low frequency of memory T_H_1* cells probably contributes to the susceptibility to mycobacterial infection observed in patients with RORγ/RORγT deficiency. As memory T_H_17 and T_H_1* cells together account for almost all *Candida*-specific CD4^+^ T cells in humans *in vivo* (Acosta-Rodriguez et al., 2007), the lower abundance of both in RORγ/RORγT deficiency probably underlies the susceptibility to *Candida* in these patients.

### A distinctive transcriptomic profile in RORγ/RORγT deficiency

We then evaluated the transcriptomic landscape of RORγ/RORγT deficiency at the single-cell level. Single-cell RNA sequencing (scRNA-seq) was performed on PBMCs from CTRLs, patients with IL-12Rβ1deficiency, and patients with RORγ/RORγT deficiency. Principal component analysis (PCA) revealed three distinct clusters corresponding to CTRLs, IL-12Rβ1deficiency, and RORγ/RORγT deficiency, respectively (**Fig. 5A**). Unbiased clustering resolved 23 unique leukocyte subsets (**Fig. 5B and 5C**). Consistent with findings from spectral flow cytometry (**Fig. 4**), patients with RORγ/RORγT deficiency had lower frequencies of naïve CD8^+^ T cells, MAIT cells, T_H_1*, and T_H_17 cells, an enrichment in T_H_1 cells, naïve B cells, and Vδ1^+^ γδ T cells, and they also displayed a trend toward lower frequencies of naïve CD4^+^ T cells (**Fig. 5C–5E**). In addition, scRNA-seq showed enriched T_H_2 cells in RORγ/RORγT deficiency (**Fig. 5D**). Differential expression was analyzed separately for each of the 23 leukocyte subsets, resulting in a combined list of 125 genes displaying significant up- or downregulation in RORγ/RORγT deficiency relative to CTRLs (**Fig. 5F** and **Table 3**). Distinctive immune gene signatures were observed in RORγ/RORγT deficiency but were restricted to T-cell compartments, including naïve CD4^+^ and CD8^+^ T cells in particular (**Fig. S4A**). Both naïve CD4^+^ and CD8^+^ T cells displayed enrichment in signatures of exhaustion and skewing toward a memory phenotype (**Fig. S4A**). A focused analysis of selected gene modules related to T-cell differentiation and activation, including naïveness, activation, inflammation, cytokine signaling, and stress, demonstrated the same loss of the naïve T-cell signature in both naïve CD4^+^ and CD8^+^ T cells from patients with RORγ/RORγT deficiency (**Fig. S4A**). We therefore focused on distinct RORγT-dependent transcriptomic profiles in naïve CD4^+^ and CD8^+^ T cells (**Fig. S4C and S4D**). In naïve CD4^+^ cells, 41 genes were upregulated relative to CTRLs in RORγ/RORγT deficiency, but not in IL-12Rβ1 deficiency (**Fig. S4C**). Notably, naïve CD4^+^ T cells from RORγ/RORγT-deficient patients overexpressed *TOX*, *TOX2*, and *IKZF2*, three transcription factors associated with the chronic activation and exhaustion of T cells (Akimova et al., 2011; Alfei et al., 2019; Neyens et al., 2023; Seo et al., 2019). In naïve CD8^+^ T cells, 18 genes were upregulated relative to CTRLs in RORγ/RORγT deficiency, but not in IL-12Rβ1deficiency (**Fig. S4D**). The overexpression of *IKZF2*, which was common to naïve CD4^+^ T cells, together with that of *GPR171* and *TAB2*, provides further support for the existence of a chronic activation and exhaustion-associated phenotype (Akimova et al., 2011; Fujiwara et al., 2021; Kanayama et al., 2004; Neyens et al., 2023). We mapped transcription factor (TF) activities in naïve CD4^+^ and CD8^+^ T cells. STAT1, STAT3, LEF1, NF-κB, E2F and SMAD TF-family activities were significantly upregulated in naïve T cells from patients with RORγ/RORγT deficiency (**Fig. 5G**). The STAT1- and IRF1-associated TF signals in RORγ/RORγT-deficient patients appeared to be driven predominantly by type I IFN/ISG rather than type II IFN (**Fig. S4E**). Enhanced NF-κB and E2F TF signals in naïve T cells suggested skewing toward T-cell activation, consistent with other analyses. However, the enrichment in LEF1 TF activity is indicative of an enhancement of progenitor-like features and of the homeostatic proliferation of naïve T cells in RORγ/RORγT deficiency, potentially providing an explanation for T-cell recovery over time in these patients (Galletti et al., 2020; Shan et al., 2021). Similarly, *RTKN2*, encoding a Rho-GTPase effector protein that promotes T-cell survival and cell-cycle progression (Bishop and Hall, 2000; Choi et al., 2024), was also overexpressed in multiple CD4^+^ and CD8^+^ T-cell subsets, including naïve CD4^+^ T cells, T_H_1, T_H_2, T_H_1*, naïve CD8^+^ T cells, CD8^+^ CM, CD8^+^ EM, and activated T cells (**Fig. 5F**). CD4^+^ and CD8^+^ T cells are, therefore, skewed toward a memory phenotype, but compensatory mechanisms involving at least LEF1 and RTKN2 may contribute to the survival and proliferation programs underlying T-cell recovery in these patients as they age.

**Figure 5.**
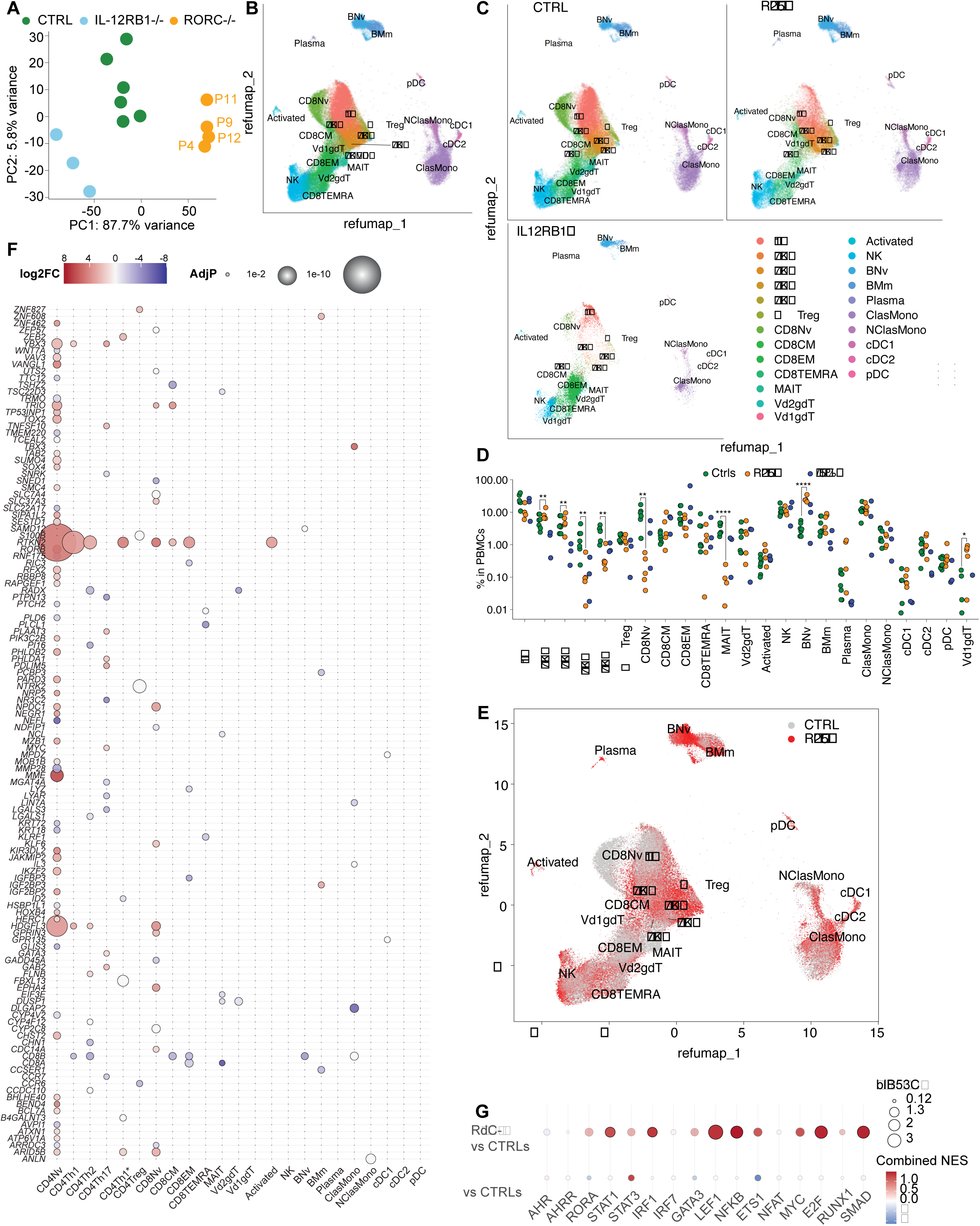
Single-cell transcriptomic profiling of immune subsets in patients with inherited RORγ/RORγT deficiency. (**A**) Principal component analysis (PCA) of scRNA-seq data from PBMCs of healthy adult controls and patients with AR IL-12Rβ1 or RORγ/RORγT deficiencies. (**B**) Unbiased clustering and annotation of major immune subsets based on combined scRNA-seq data from all samples. (**C**) Distinct clustering of immune subsets from PBMCs of controls and patients. (**D**) Relative abundance of immune subsets in healthy adult controls and patients with AR IL-12Rβ1 or RORγ/RORγT deficiencies. (**E**) Differential enrichment or depletion of immune subsets in RORγ/RORγT-deficient patients relative to healthy adult controls. (**F**) Bubble plot showing all genes significantly enriched or depleted (*p*-value <= 0.01) in each immune subset in patients with RORγ/RORγT deficiency relative to controls. Bubble size reflects the adjusted *p*-value; color indicates the fold-change difference. The absence of a bubble indicates an adjusted *p*-value > 0.01 for the subset concerned. (**G**) TF-family enrichment in aggregated naïve CD4^+^ and CD8^+^ T cells from RORγ/RORγT- or IL-12Rβ1-deficient patients relative controls. Color indicates normalized enrichment score (NES); size indicates −log10 false-discovery rates.

**Table 3:**
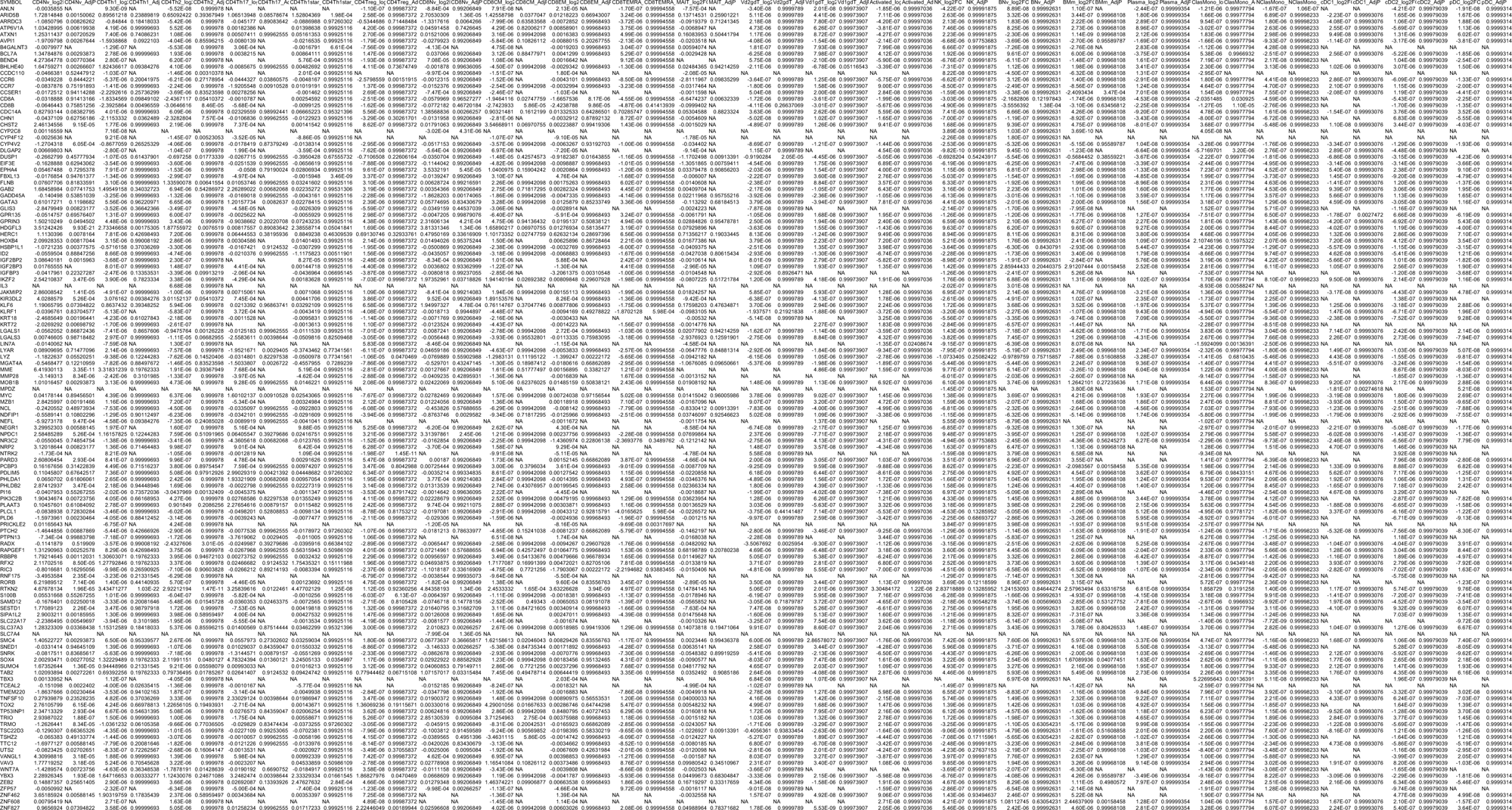

### Altered T helper cell effector function in patients with RORγ/RORγT deficiency

We then assessed the effector function of CD4^+^ T cells from patients P9 and P11, focusing on both naïve and memory subsets. Naïve CD4^+^ T cells were activated under T_H_0-, T_H_1-, T_H_2-, and T_H_17-polarizing conditions. Differentiating CD4^+^ T cells from both P9 and P11 displayed low levels of IFN-γ and TNF production under T_H_0-polarizing conditions (**Fig. 6A and 6B**). Levels of IFN-γ and TNF production by P9 and P11 under T_H_1-polarizing conditions were also lower than those in most healthy controls (**Fig. 6A and 6B**). Consistent with the absence of T_H_17 cells *ex vivo*, naïve CD4^+^ T cells from patients failed to produce IL-17A and IL-17F upon T_H_17 differentiation (**Fig. 6C and 6D**). Like activated naïve CD4^+^ T cells, memory CD4^+^ T cells from P9 and P11 subjected to activation under T_H_0- and T_H_1-polarizing conditions also displayed a significant impairment of IFN-γ production and a trend toward lower than normal levels of TNF production (**Fig. 6E and 6F**). These results, together with the data for naïve CD4^+^ T cells (**Fig. 6A**), suggest that the impairment of IFN-γ secretion in RORγ/RORγT deficiency stems from both the smaller numbers of IFN-γ-producing T_H_1* cells and a cell-intrinsic defect of a RORγT-dependent mechanism regulating IFN-γ production. Memory CD4^+^ T cells from P9 and P11 failed to produce any IL-17A or IL-17F under T_H_0- or T_H_17-polarizing conditions (**Fig. 6G and 6H**). Interestingly, although naïve CD4^+^ T cells from P9 and P11 displayed no abnormal production of T_H_2 cytokines (data not shown), memory CD4^+^ T cells from P9 and P11 produced abnormally high levels of IL-5 (**Fig. 6I**), suggesting that the high levels of T_H_2 cytokine production in RORγ/RORγT deficiency are driven principally by memory CD4^+^ T cells. Overall, our data demonstrate that RORγ/RORγT promotes the production of IFN-γ, TNF, and IL-17A/F via a CD4^+^ T cell-intrinsic mechanism. The absence of IL-17A/F-mediated immunity underlies susceptibility to mucocutaneous *Candida* infection in patients with inherited RORγ/RORγT deficiency, consistent with findings for other IEIs affecting IL-17A/F signaling (Puel, 2020; Tangye and Puel, 2023).

**Figure 6.**
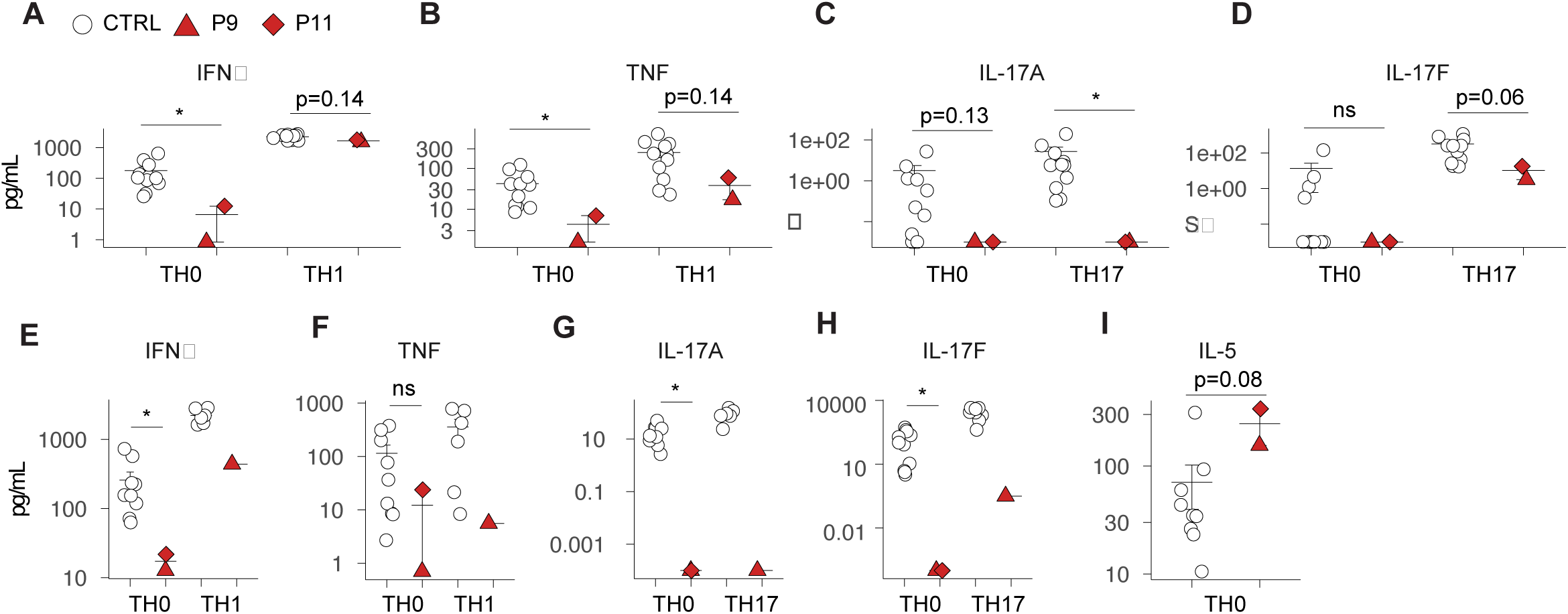
Assessment of the effector function of CD4^+^ T-cell subsets. (**A - D**) Production of IFN-γ (A), TNF (B), IL-17A (C), IL-17F (D), by naive CD4^+^ T cells from healthy controls and RORγ/RORγT-deficient patients cultured under T_H_0-, T_H_1-, and T_H_17-polarizing conditions. (**E – I**) Production of IFN-γ (E), TNF (F), IL-17A (G), IL-17F (H), and IL-5 (I) by memory CD4^+^ T cells stimulated under T_H_0-, T_H_1-, and T_H_17-polarizing conditions. Statistical significance was determined using two-sided Wilcoxon rank-sum tests with Benjamini-Hochberg correction for multiple comparisons. All experimental data were verified in at least 2 independent experiments.

### Impaired antimycobacterial immunity in patients with RORγ/RORγT deficiency

We investigated the leukocyte subsets responsible for RORγ/RORγT-dependent IFN-γ production upon mycobacterial infection to elucidate the molecular and cellular basis of mycobacterial diseases in patients with inherited RORγ/RORγT deficiency. PBMCs from CTRLs, patients with IL-12Rβ1 deficiency, and patients with RORγ/RORγT deficiency, including P9, P11, P12, and the previously published P4, were infected with live BCG *in vitro*, in the presence or absence of exogenous IFN-γ, IL-12 or IL-23, cytokines with well-established roles in antimycobacterial immunity through the promotion of IFN-γ production by various immune cells (Boisson-Dupuis, 2020; Bustamante, 2020; Casanova and Abel, 2022; Kerner et al., 2020; Martínez-Barricarte et al., 2018; Ogishi et al., 2022; Philippot et al., 2023). Leukocytes from CTRLs and patients with RORγ/RORγT deficiency produced large amounts of IFN-γ upon stimulation with IL-12 or IL-23 (**Fig. 7A**) whereas those from patients with IL-12Rβ1 deficiency did not. However, RORγ/RORγT-deficient leukocytes produced less IFN-γ than leukocytes from CTRLs upon stimulation with IL-23 (**Fig. 7A**). Following infection with BCG *in vitro*, leukocytes produced very high levels of multiple inflammatory cytokines. However, leukocytes from RORγ/RORγT-deficient patients produced less IFN-γ and TNF than CTRLs during infection, regardless of the presence or absence of IL-12 or IL-23 (**Fig. 7A and 7B**). By contrast, the production of IL-6, IL-8, GM-CSF, and IL-1β was similar in CTRLs and patients with RORγ/RORγT deficiency (**Fig. 7C and Fig. S5A**). Thus, the production of IFN-γ and TNF — two key antimycobacterial effector cytokines in humans (Arias et al., 2024; Kerner et al., 2020) — is selectively impaired during mycobacterial infection in individuals with RORγ/RORγT deficiency.

**Figure 7.**
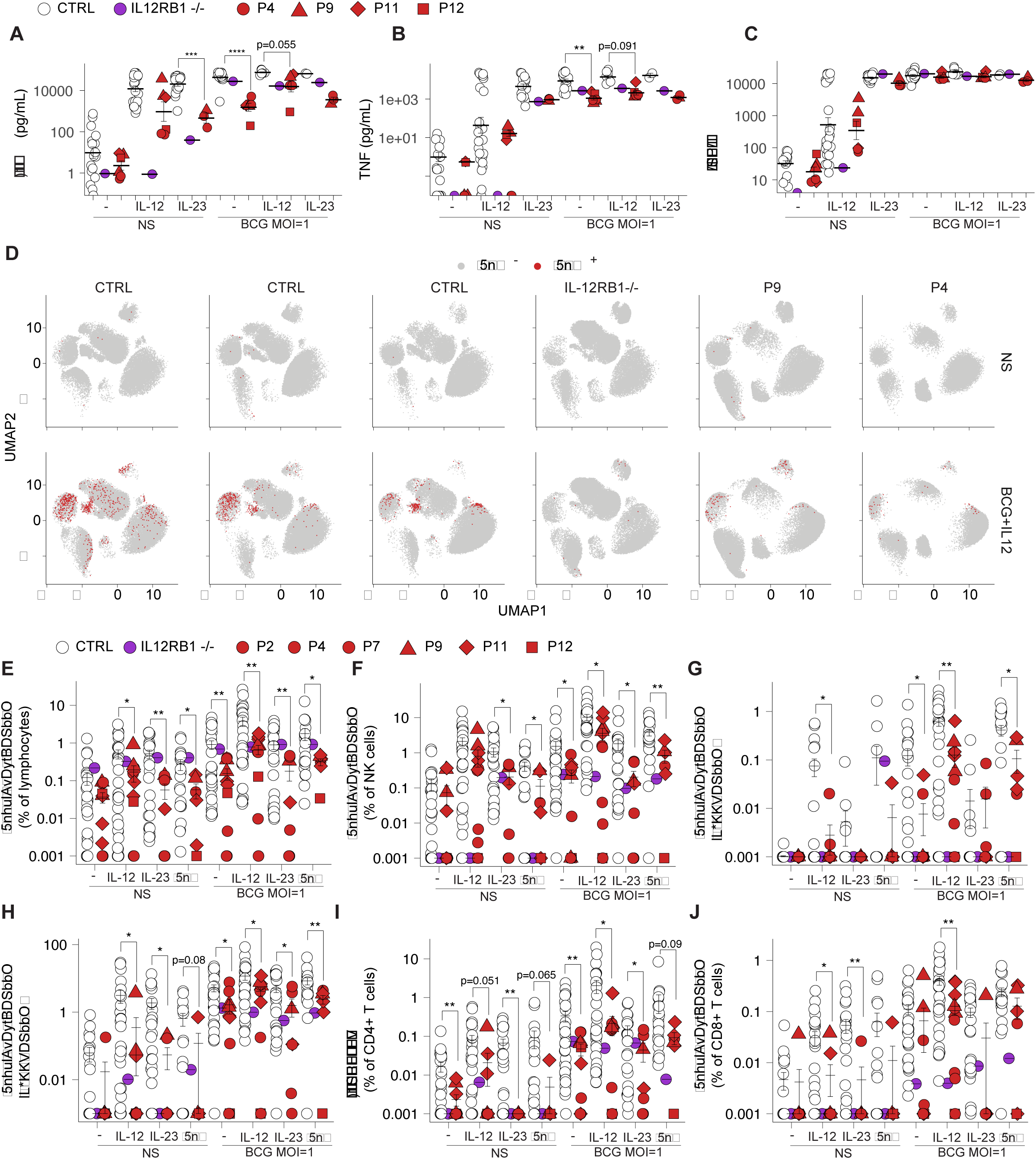
Analysis of cytokines governing antimycobacterial immunity during mycobacterial infection. (**A - C**) Production of IFN-γ (A), TNF (B), and IL-6 (C) by PBMCs with and without live BCG infection, in the presence or absence of exogenous IL-12 or IL-23. (**D**) UMAP visualization of IFN-γ-producing leukocyte subsets in PBMCs from healthy controls, patients with IL-12Rβ1 deficiency, and patients with RORγ/RORγT deficiency. (**E - J**) Frequencies of IFN-γ-producing cells among total lymphocytes (E), and within specific immune subsets including NK cells (F), Vδ1^+^ γδ T cells (G), Vδ2^+^ γδ T cells (H), CD4^+^ T cells (I), and CD8^+^ T cells (J), following *in vitro* infection with live BCG, in the presence or absence of exogenous IL-12, IL-23, or IFN-γ. All experimental data were verified in at least 2 independent experiments.

### Poor innate and innate-like adaptive antimycobacterial immunity

For identification of the lymphoid subsets responsible for RORγ/RORγT-dependent antimycobacterial immunity, we assessed the levels of both lineage-specific surface markers and intracellular cytokines by spectral flow cytometry during acute BCG infection *in vitro*, in the presence or absence of IL-12, IL-23, and IFN-γ. Major lymphoid subsets were identified by unbiased clustering based on the distinctive expression patterns of lineage-determining surface markers (**Fig. S5B**). The production of both IFN-γ and TNF increased in response to BCG infection in PBMCs from healthy donors, especially in the presence of exogenous IL-12, but lymphocytes from patients with RORγ/RORγT deficiency had markedly lower levels of IFN-γ production regardless of exogenous cytokine supplementation (**Fig. 7D, E, and Fig. S5C**). We analyzed each leukocyte subset separately, to identify the specific lymphocyte subsets responsible for differential IFN-γ production. NK cells from patients with RORγ/RORγT deficiency — including both CD56^bright^ and CD16^+^CD56^dim^ NK cells — had lower intracellular IFN-γ and TNF levels during BCG infection, regardless of the exogenous cytokine conditions (**Fig. 7F and Fig. S5D–S5F**). The innate-like adaptive lymphocyte population was depleted of MAIT and iNKT cells (**Fig. S5B**). Despite the enrichment in Vδ1^+^ γδ T cells observed in patients with RORγ/RORγT deficiency, these cells produced markedly low levels of IFN-γ and TNF during BCG infection in the presence or absence of IL-12 (**Fig. 7G and Fig. S5G**). Vδ2^+^ γδ T cells, which normally display robust T-bet-dependent IFN-γ and TNF production in response to BCG infection (Yang et al., 2020), were also functionally impaired in patients with RORγ/RORγT deficiency, producing significantly smaller amounts of IFN-γ and TNF than CTRL cells (**Fig. 7H and Fig. S5H**). Thus, the innate and innate-like adaptive lymphocytes of patients with RORγ/RORγT deficiency displayed a marked depletion of MAIT and iNKT cells, and their NK cells, Vδ1^+^, and Vδ2^+^ γδ T cells displayed functional impairment during acute BCG infection, despite their preserved development. MAIT, iNKT, NK, and Vδ2^+^ γδ T cells are also adversely affected — in terms of their numbers, their IFN-γ-producing capacity, or both — in AR T-bet deficiency (Yang et al., 2020), suggesting that the mechanisms affecting innate and innate-like adaptive lymphocytes may be common to inherited T-bet and RORγ/RORγT deficiencies.

### Markedly lower levels of IFN-γ-producing αβ T cells in patients with RORγ/RORγT deficiency

We then investigated the role of αβ T cells in antimycobacterial immunity during BCG infection. In CTRLs, exogenous IL-23 increased the frequency of IFN-γ-producing CD4^+^ T cells in the absence of BCG infection. By contrast, CD4^+^ T cells from RORγ/RORγT-deficient patients failed to respond to IL-23 stimulation (**Fig. 7I**). During acute BCG infection, the frequency of IFN-γ-producing CD4^+^ T cells was consistently lower for cells from patients with RORγ/RORγT deficiency than for control cells, regardless of the exogenous cytokines applied, with the largest difference observed in the presence of exogenous IL-12 (**Fig. 7I**). However, TNF production by CD4^+^ T cells was less strongly affected (**Fig. S5I**). Circulating mature CD8^+^ T-cell counts were markedly lower in patients with RORγ/RORγT deficiency (**Fig. 4 and Fig. 5**), but the residual CD8^+^ T cells also produced smaller amounts of IFN-γ upon BCG infection in the presence of IL-12 (**Fig. 7J**). However, TNF production by CD8^+^ T cells during BCG infection was preserved (**Fig. S5J**). Thus, αβ T cells — both CD4^+^ T and CD8^+^ T cells — displayed impaired IFN-γ production, contributing to the weaker antimycobacterial immunity in patients with RORγ/RORγT deficiency. The observed impairment probably results from both the smaller numbers of *Mycobacterium*-specific precursors and intrinsic defects of RORγT-governed IFN-γ production.

### IFN-γ production upon prolonged stimulation with *Mycobacterium* is mediated by RORγT

We investigated the possible intrinsic role of RORγT in regulating IFN-γ production during BCG infection by stimulating PBMCs from BCG-vaccinated CTRLs, patients with IL-12Rβ1 deficiency, and patients with RORγ/RORγT deficiency, including P4 and P9, with purified protein derivative (PPD, or tuberculin) or lysates derived from BCG (BCG-L), for eight days. Actively proliferating cells, characterized as CellTrace^−^Ki67^+^ cells, were selected for comprehensive gating based on their surface marker expression and analysis of their intracellular cytokine production. αβ T cells, including antigen-specific CD4^+^ T cells, proliferated robustly in response to PPD, whereas Vδ2^+^ γδ T cells, which recognize small organic phosphate antigens (phosphoantigens, or pAgs) in BCG-L (Herrmann et al., 2020; Rhodes et al., 2016) were the dominant responders during BCG-L stimulation. Following stimulation with either BCG-L or PPD, proliferating *Mycobacterium*-specific lymphocytes produced IFN-γ slowly but steadily, with strong amplification following re-stimulation with P/I (**Fig. 8A and 8B**). PPD-specific lymphocytes (**Fig. 8C**), particularly CD4^+^ T cells (**Fig. 8D**), but not NK, or Vδ2^+^ γδ T cells (**Fig. 8E and 8F**) from patients with RORγ/RORγT deficiency displayed slightly lower levels of IFN-γ production both with and without P/I re-stimulation. By contrast, proliferating BCG-L-specific lymphocytes, which were mostly Vδ2^+^ γδ T cells, did not display significantly lower levels of IFN-γ production (**Fig. 8B**). These findings suggest that RORγT is required for robust antimycobacterial responses during acute infection, but that this response becomes attenuated and less RORγT-dependent in innate immune and innate-like adaptive T cells subjected to prolonged stimulation. A small subset of PPD-reactive CD4^+^ T cells from CTRLs also produced IL-17A and IL-17F (**Fig. 8G and 8H**) and displayed high levels of RORγT expression (**Fig. 8I**). These findings confirm that *Mycobacterium*-specific T cells are derived from T_H_1* cells (Acosta-Rodriguez et al., 2007). However, cells producing IL-17A or IL-17F cells were absent from the PPD-reactive CD4^+^ T cells of patients with inherited RORγ/RORγT deficiency (**Fig. 8I**). Thus, RORγT is essential not only for the correct development of *Mycobacteria*-specific T_H_1* cells but also for the intrinsic induction of IFN-γ production in these cells.

**Figure 8.**
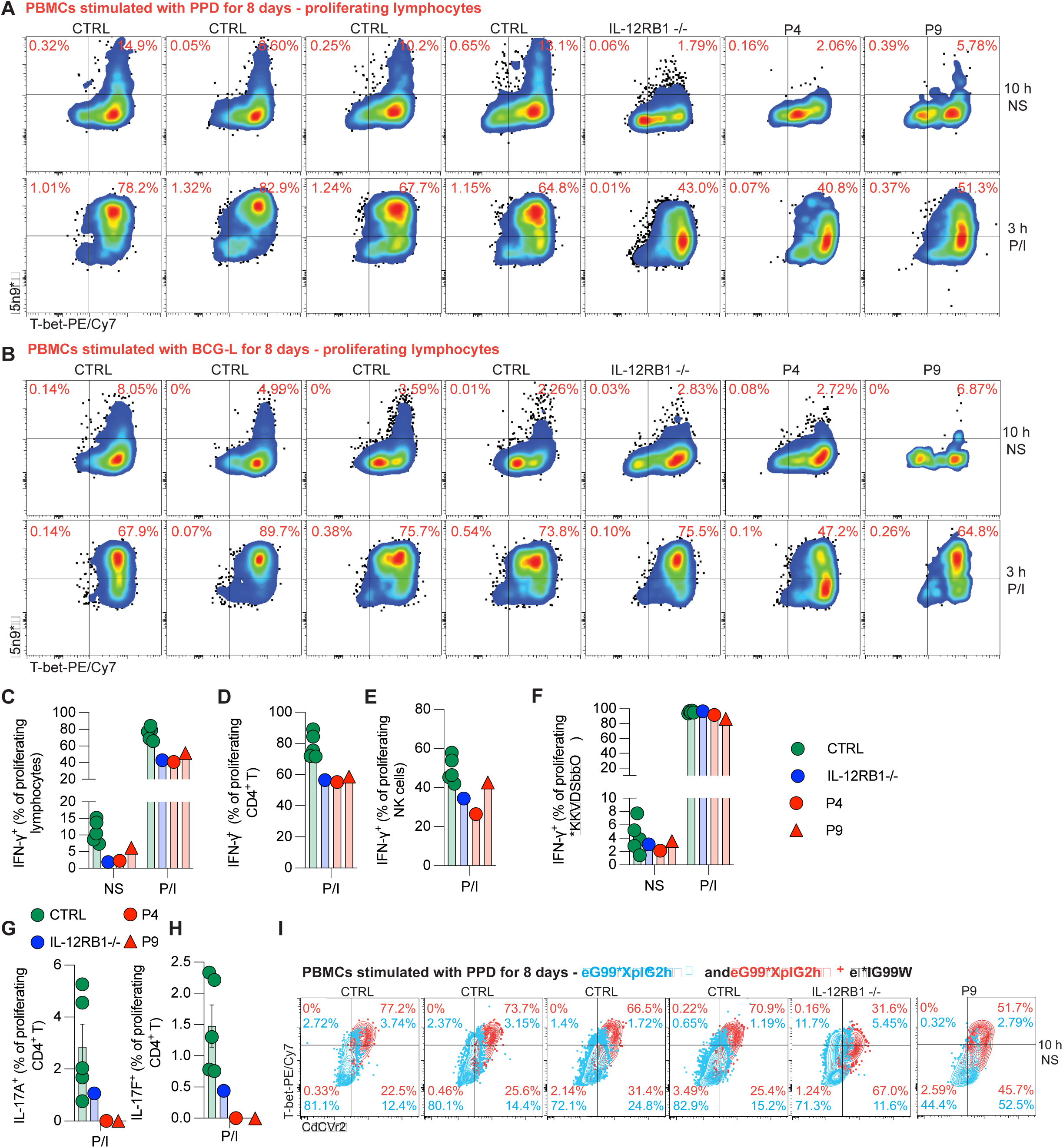
Antimycobacterial immunity in *Mycobacterium*-specific lymphocytes. (**A** and **B**) Dot plots showing IFN-γ production and T-bet expression in proliferating *Mycobacterium*-specific lymphocytes (CellTrace^−^Ki67^+^) from BCG-vaccinated healthy controls, and patients with IL-12Rβ1 or RORγ/RORγT deficiencies, following stimulation with purified protein derivative (PPD) (A) or with BCG-L (B). (**C - F**) Frequencies of intracellular IFN-γ-producing cells among PPD-specific proliferating total lymphocytes (C), CD4^+^ T cells (D), NK cells (E), and Vδ2^+^ γδ T cells (F). (**G and H**) Frequencies of IL-17A- (G) and IL-17F-producing (H) cells among PPD-specific proliferating CD4^+^ T cells. (**I**) Comparison of intracellular RORγT and T-bet expression levels in PPD-specific proliferating versus non-proliferating CD4^+^ T cells from healthy controls and patients. All experimental data were verified in at least 2 independent experiments.

## Discussion

Here, we expand the genotypic spectrum of inherited RORγ/RORγT deficiency by identifying five new disease-causing variants in the homozygous state in five unrelated individuals. In total, eight disease-causing variants have been described, affecting all three functional domains of RORγT: two missense variants in the DBD (S17L and V24F), two truncating variants in the hinge domain (R77* and Y165*), and one missense plus three truncating variants in the LBD (Q308*, K360I, T400Nfs*56, and Q420*) (Zou et al., 2022). All variants identified to date result in a complete LOF for transcriptional activity. The discovery of one of these variants, K360I, is particularly noteworthy. The LBD adopts a transcriptionally active conformation even in the absence of ligand binding (Zou et al., 2022). The activity of this domain is modulated by diverse endogenous and synthetic ligands, including cholesterol-related intermediates, oxysterols, terpenoids, polyketides, and cardiac glycosides (Jetten and Cook, 2020; Ladurner et al., 2021). The LBD is, therefore, a compelling target for inverse agonists for the treatment of autoimmune and immune dysregulation disorders (Jetten and Cook, 2020). However, no RORγ/RORγT-targeting agents have yet advanced to phase III clinical trials due to efficacy concerns (Zeng et al., 2023). In this context, the K360I variant provides unique insight. Located within the LBD, K360I completely abolishes the transcriptional activity of RORγT. These findings reveal an essential role for the K360 residue in RORγT function, positioning it as a promising target for the design of more selective and effective inverse agonists.

We further expanded the clinical spectrum of inherited RORγ/RORγT deficiency by detailing its clinical manifestations, natural course, and prognosis. Remarkably, inherited complete RORγ/RORγT deficiency underlies consistent clinical phenotypes across seven ancestries (Palestinian, Chilean, Saudi Arabian, Afghan, Sri Lankan, Indian, and Iranian) and in diverse environments from four different regions: Australia, South Asia, the Middle East, and South America. With the exception of P11, who benefited from early prophylaxis, all patients presented with severe mycobacterial disease. Nine patients presented with BCG-osis, one with *M. fortuitum* complex pneumonia, one with TB lymphadenitis, and one with disseminated tuberculosis followed, after nine TB-free years, by osteoarticular TB. Despite vaccination with BCG, neither P5 nor P10 developed BCG-itis or BCG-osis; instead, both presented with tuberculosis. No recurrence of NTM infection was observed, whereas TB recurred in P10, with both episodes in this patient being extrapulmonary. The natural course of inherited RORγT deficiency therefore suggests that TB recurrence, particularly in regions in which this disease is endemic, is a genuine risk, consistent with the much greater virulence of *Mtb* than of NTM (Casanova and Abel, 2022). Deficiencies of T-bet and RORγT, two transcription factors governing commitment to the T_H_1 and T_H_17 lineages (Ivanov et al., 2006; Okada et al., 2015; Szabo et al., 2000; Yang et al., 2020), respectively, lead to different but overlapping mechanisms of mycobacterial susceptibility (Ogishi et al., 2023b; Okada et al., 2015; Yang et al., 2020). In inherited T-bet deficiency, susceptibility is due to the impaired development of NK, MAIT, iNKT, and Vδ2^+^ γδ T cells, and the production of much smaller amounts of IFN-γ by these subsets, despite preserved T_H_1* responses (Yang et al., 2020). By contrast, RORγ/RORγT deficiency causes a near-complete depletion of MAIT and iNKT cells, a partial depletion of CD8^+^ T and CD4^+^ T cells, including T_H_1* cells, and impaired IFN-γ production by these subsets and by Vδ1^+^, Vδ2^+^ γδ T, and NK cells.

These findings expand those of our previous report (Okada et al., 2015). MAIT and iNKT cells recognize *Mycobacterium*-derived metabolites via MR1 and CD1d, respectively, and are among the most robust early responders to BCG or *Mtb* (Sada-Ovalle et al., 2008; Vorkas et al., 2018; Yang et al., 2020). Critically, there is a complete or partial depletion of these cells in inherited RORγ/RORγT and T-bet deficiencies. However, the loss of each cell type alone is insufficient to confer mycobacterial susceptibility, as demonstrated by the absence of mycobacterial disease in patients with AR MR1 deficiency and X-linked lymphoproliferative disease 1 (XLP1) (Howson et al., 2020; Tangye and Latour, 2020), who lack MAIT and iNKT cells, respectively. By contrast, inherited TCRα constant chain deficiency, which depletes both MAIT and iNKT cells (Rawat et al., 2021), confers a predisposition to mycobacterial disease, with 25% of affected patients developing BCG-osis (Materna et al., 2025). The loss of IFN-γ-producing subsets or IFN-γ-responsiveness, in addition to MAIT and iNKT cells, appears to be crucial for the higher penetrance of mycobacterial disease. For example, the impairment of T_H_1* cells in inherited deficiencies of SPPL2A or LY9 in humans contributes to mycobacterial disease (Bustamante, 2020; Kong et al., 2018; Ogishi et al., 2025). Thus, impaired IFN-γ production by Vδ2^+^ γδ T and NK cells in inherited T-bet deficiency, and the overall decrease in CD4^+^ T-cell counts, particularly for T_H_1* and CD8^+^ T cells, together with broad IFN-γ production defects in CD4^+^, CD8^+^ αβ T, Vδ1^+^, Vδ2^+^ γδ T, and NK cells in inherited RORγ/RORγT deficiency, probably result in a tipping point leading to mycobacterial susceptibility.

With the exception of P11, who received early fluconazole prophylaxis, all but one patient (P3, who was lost to follow-up early), developed CMC of variable severity, directly attributable to the abolition of IL-17A and IL-17F production by CD4^+^ T cells and probably to the depletion of IL-17-producing MAIT cells. There is pathway homogeneity between this mechanism and those underlying inborn errors of IL-17 cytokines or their receptors (Okada et al., 2016; Puel, 2020; Tangye and Puel, 2023). However, several features are notable. First, the clinical course of CMC in RORγ/RORγT-deficient patients is typically mild, often responsive to topical and systemic triazole antifungal drugs, with sustained remission even in the absence of prophylaxis or poor compliance with prophylaxis. This, together with the incomplete penetrance of this deficiency, resembles the clinical phenotype of AD IL-17F, AR IL-23R, and AR IL-17RC deficiencies, rather than the more severe, refractory CMC, and the often highly penetrant staphylococcal skin infections seen in AR IL-17RA or AR ACT1 deficiencies (Lévy et al., 2016; Philippot et al., 2023; Puel, 2020). These findings suggest that either some residual IL-17A/F-producing cells persist in patients with inherited RORγ/RORγT deficiency, or that the production of certain IL-17 cytokines may occur independently of RORγ/RORγT, or both. Notably, T_H_17 memory CD4^+^ T cells were present at low levels but were not entirely absent in these patients. Other known IL-17-producing cells, including tissue-resident T cells and ILC3s, were too rare for evaluation in peripheral blood. Furthermore, the role of RORγ/RORγT in the production of non-IL-17A/F IL-17 cytokines, such as IL-17E/IL-25 and IL-17C, remains unclear. IL-17C is dispensable for anti-*Candida* immunity in mice (Conti et al., 2015), but IL-25 has been shown to promote antifungal responses by inducing IL-17A/F production in a specific ILC subset in mice (Huang et al., 2015; Lévy et al., 2016). These findings suggest that other cytokines, including but not limited to IL-25, may confer partial and redundant protection against CMC and *S*. *aureus* in the absence of RORγ/RORγT.

Several less common but clinically meaningful phenotypes were observed in patients with RORγ/RORγT deficiency. T_H_2 polarization is enhanced in both inherited T-bet and RORγ/RORγT deficiencies. However, only one of the 12 RORγ/RORγT-deficient patients displayed T_H_2 diathesis, contrasting with the single T-bet-deficient patient who had refractory T_H_2-disease (Yang et al., 2020; Yang et al., 2021). Thus, although T_H_2 skewing is immunologically evident from the increase in T_H_2 cytokine levels without a concomitant increase in IgE levels, it displays only weak clinical penetrance in RORγ/RORγT deficiency. Two patients (P7 and P10) experienced cutaneous warts due to HPV infection, perhaps associated with their limited usage of distal *TRAV* and *TRAJ* segments resulting in T-cell lymphopenia. This mechanism is reminiscent of the weakly penetrant HPV susceptibility observed in ~ 25% of patients with inherited TCRα constant chain deficiency (Materna et al., 2025; Rawat et al., 2021), ~ 27% patients with idiopathic CD4 lymphopenia (Yarmohammadi and Cunningham-Rundles, 2017), and a sizeable proportion of patients with protein-losing enteropathy (Vignes and Bellanger, 2008). Alternatively, RORγT deficiency may compromise the development of skin-homing T cells, tissue-resident ILCs or T cells contributing to local anti-HPV immunity. Nevertheless, the cutaneous warts observed in these two patients appeared to be self-limiting. The narrow infectious phenotypes observed in patients with inherited RORγ/RORγT deficiency serve as a natural model for anticipating and mitigating the potential adverse effects of RORγT antagonists in clinical trials. T-cell ALL was observed in P7. Notably, both germline and adult-induced conditional RORγT knockout mice have been reported to develop thymic T-cell lymphoma (Liljevald et al., 2016; Ueda et al., 2002). These tumors may arise from a CD4^−^CD8^−^ DN T-cell clone (Ueda et al., 2002). Long-term follow-up of our patients will be required to clarify the impact of RORγ/RORγT deficiency on the risk of T-cell malignancy in humans.

Importantly, TRECs were undetectable or present at abnormally low levels in all three patients tested, a hallmark of profound T-cell lymphopenia, including SCID or leaky SCID (Dvorak et al., 2023; Shearer et al., 2014). The T-cell counts in all 12 patients were within the leaky SCID range but markedly above the classic SCID level (Dvorak et al., 2023). The naïve T-cell population expanded with age, and no severe opportunistic infections with microbes other than mycobacteria were observed. For the four patients for whom vaccination records were available, all four tolerated live-attenuated MMR or varicella vaccination. The absence of TRECs probably reflects the defective TCRα rearrangement (Berland et al., 2019). Thus, a diagnosis of inherited RORγ/RORγT deficiency should be considered in infants in whom TRECs are absent or present at only low levels during newborn screening. Furthermore, restricted TCRα usage, along with significant expansion of the DN T-cell and Vδ1^+^ γδ T-cell populations, may be used to distinguish between this condition and leaky SCID. Finally, all but one of the RORγ/RORγT-deficient patients (aged 2 to 20 years) have survived without hematopoietic stem-cell transplantation (HSCT), and those receiving appropriate antimycobacterial and antifungal prophylaxis remain clinically stable. These findings suggest that HSCT is probably not necessary for patients with inherited RORγ/RORγT deficiency.

## METHODS

### Human subjects

Studies involving data and samples from patients and healthy volunteers were approved by the institutional review and privacy boards (IRB) and local ethics committees, in accordance with the Declaration of Helsinki. Experiments were conducted in the United States, Australia, and France in compliance with local regulations and with approval from the Institutional Review Board (IRB) of The Rockefeller University (protocol no. JCA-0700), INSERM (protocol no. C10-13), and the Sydney Local Health District RPAH Zone Human Research Ethics Committee and Research Governance Office, Royal Prince Alfred Hospital, Camperdown, NSW, Australia (protocols X20-0177 & 2020/ETH0098; X16-0210/LNR/16/RPAH/257; and X16-0210 & 2019/ETH06359).

Written informed consent was obtained from all patients and their family members in their countries of residence, in accordance with local regulations and IRB approvals. Patients P8 and P12 were recruited under a protocol approved by the Ethics Committee of Tehran University of Medical Sciences, Iran (protocol no. IR.TUMS.CHMC.REC.1397.004). Patient P9 was recruited under a protocol approved by the Human Research Ethics Committee of the Royal Children’s Hospital, Melbourne, Australia (HREC 3316). Patient P10 was recruited under protocol C10-13 approved by INSERM. Patient P11 was recruited under an IRB approved protocol at the University of Miyazaki, Japan. Healthy controls were recruited locally at The Rockefeller University, New York, USA (protocol no. JCA-0709) and the Imagine Institute, Paris, France, in accordance with local regulations. Buffy coats from healthy donors were obtained from the Australian Red Cross Blood Service. Written informed consent was obtained from all healthy volunteers enrolled in this study.

### Extended case report

Extended case reports for P8–P12 are available from corresponding authors upon request.

### Whole-exome sequencing

The method used for whole-exome sequencing (WES) has been described elsewhere (Arias et al., 2024). Genomic DNA from P8 - P12 and their relatives was used for WES. Exome capture was performed with the SureSelect Human All Exon V4+UTR and SureSelect Human All Exon V6 kits (Agilent Technologies). Paired-end sequencing was performed on a HiSeq 2000 sequencer (Illumina) generating 100-base reads. We used the Genome Analysis Software Kit (GATK) (v.3.4-46) best-practice pipeline to analyze our WES data. Reads were aligned with the human reference genome GRCh38, with BWA. PCR duplicates were removed with Picard tools v.3.1.1 (https://picard.sourceforge.net/). The GATK base quality-score recalibrator was used to correct sequencing artifacts. The GATK HaplotypeCaller v.4.1.4.1 was used to identify variant calls. Variants were annotated with SnpEff v.4.5 (https://snpeff.sourceforge.net/). Homozygosity rates were estimated from the patients’ genomic DNA, as previously described (Fareed and Afzal, 2017). Parametric multipoint linkage analysis was performed on the WES data with MERLIN v.1.1.2, assuming AR inheritance with complete penetrance and a damaging allele frequency of 1 × 10^−4^. Allele frequencies were estimated for 72,817 SNPs with the gnomAD v.2.1.1 American population. Markers were clustered with an *r*^2^ threshold (--rsq parameter) of 0.4. Of note, sharing of raw WES data was not permitted under the approved IRB protocols.

### Sanger sequencing

Genomic DNA was extracted from whole-blood samples from the patients and their healthy relatives. PCR products were sequenced with the BigDye Terminator Cycle Sequencing Kit (Applied Biosystems). Sequencing products were purified with Sephadex G-50 Superfine Resin (GE Healthcare). Sequences were determined with an ABI 3730 DNA Analyzer (Applied Biosystems). Sequencing spectrum data were analyzed with Geneious software (https://www.geneious.com).

### Immunoblotting

Mutant RORγ- and RORγT-expressing vectors were generated by site-directed mutagenesis with the pCMV6-*RORC*-Myc-DDK vector (Origene). Plasmids were used to transfect HEK293T cells in the presence of Lipofectamine 2000 reagent (Thermo Fisher Scientific), according to the manufacturer’s protocol. Nuclear and cytoplasmic extracts were prepared with NE-PER Nuclear Extraction Reagent (G Biosciences), 24 hours post transfection. Immunoblotting was performed with antibodies targeting RORγ (13205-1-AP: Proteintech; sc-365476: Santa Cruz; 29910-1-AP: Proteintech), lamin A/C (E-1 sc-376248: Santa Cruz), α-tubulin (HRP-66031: Proteintech), and vinculin (sc-73614: Santa Cruz).

### Reverse transcription-quantitative PCR (RT-qPCR)

For RT-qPCR analysis, total RNA was extracted from transfected HEK293T cells with the RNeasy Plus Micro Kit. TaqMan Gene Expression Assays were performed according to the manufacturer’s protocol, with *RORC* (Hs01076122) and Hu *GUS* probes.

### Retrovirus production and transduction of T-cell lines

The pLZRS-IRES-ΔNGFR (empty vector) and WT or mutant pLZRS-IRES-RORCiso1/2-ΔNGFR retroviral vectors, both containing a puromycin resistance cassette, were generated as previously described. ΔNGFR ORF encodes a truncated nerve growth factor receptor (NGFR/CD271) that is unable to signal and was used as a cell-surface marker of successful transduction. We transfected HEK293T cells with 1.5 µg pLZRS-IRES-ΔNGFR vector (empty, WT, or mutant Iso 1/Iso 2 RORC) together with Gag/Pol and VSV-G packaging plasmids, with X-tremeGENE 9 (Roche) according to the manufacturer’s instructions. The medium was replaced after six hours of incubation. At 60 hours post-transfection, the supernatant was collected and retroviral particles were concentrated with a Retro-X concentrator (Clontech) used in accordance with the manufacturer’s protocol.

For transduction, 0.2 million HuT78 cells expressing WT or mutant multimerized RORE were spinoculated with retrovirus-containing supernatant in a total volume of 200 µL for 2 hours. The cells were then incubated for 48 hours in the same plate, before splitting. After 10 days of culture, transduced cells were purified by MACS with a magnetic bead-conjugated anti-NGFR antibody (Miltenyi Biotec) according to the manufacturer’s instructions.

### Luciferase reporter assay

Mutant RORγ- and RORγT-expressing vectors were generated by site-directed mutagenesis with the pLZRS-IRES-deltaNFGR plasmid (Origene). These plasmids were used for retrovirus production and were then used to transduce HuT78 luciferase reporter cell lines expressing WT or mutant multimerized RORE. We then isolated stably transduced cells expressing the bicistronic CD271 surface reporter by MACS. CD271^+^ cell count was determined by flow cytometry with a PE-anti-CD271 antibody (BD Pharmingen).

The transcriptional activity of WT or mutant RORγ and RORγT was assessed in a stimulation assay. We first coated 96-well flat-bottomed plates overnight with 10 µg/mL anti-CD3 antibody (Bio X Cell BE0001-2) or left them uncoated. The next day, the plates were seeded with 200,000 CD271^+^ cells per well. Ionomycin (4 ng/mL) was added to the samples in anti-CD3 antibody-coated wells. After 8 hours of incubation, luciferase reagent (Dual-Glo Promega) was added, the plates were incubated for a further 20 minutes and luciferase activity was measured.

### Electrophoretic mobility shift assay

The mutant RORγ or RORγT expression plasmids were generated by site-directed mutagenesis on the pCMV6-RORC vector with or without a Myc-DDK tag (Origene). They were used to transfect HEK293T cells, in the presence of Lipofectamine LTX (Life Technologies), according to the manufacturer’s protocol. The transfectants were incubated with or without PMA (25 ng/mL) and ionomycin (0.1μ M) for 4 hours and were subjected to immunoblot analysis (with the Myc-DDK tag) or nuclear extraction for EMSA (without the Myc-DDK tag). Nuclear and cytoplasmic extracts were prepared with NE-PER Nuclear and Cytoplasmic Extraction Reagents (Thermo Fisher Scientific), used according to the manufacturer’s protocol. Immunoblotting was performed with antibodies directed against amino acids 131-320 of RORγ (H-190: Santa Cruz), DDK (4C5: Origene), lamin A/C (H-110: Santa Cruz), β-tubulin (T5293: Sigma-Aldrich), β-actin (A5316: Sigma-Aldrich) and GAPDH (G-9: Santa Cruz). EMSA was performed as previously described (*46*). We incubated 20 μg nuclear extract for 30 minutes with ^32^P-labeled (a-dATP) RORE-1 probe (5’-gat cAT TCT TCT ATG ACC TCA TTG GGG G-3’) or RORE-2 probe (5’-gat cAC CAA AAT GGT GTC ACC CCT GAA C-3’), designed on the basis of the conserved sequences of the *IL17A* promoter region (*48*).

### RT-qPCR for *IL17A*

Jurkat cells were electroporated with WT or mutant RORC plasmids with the Neon NxT Electroporation System (Thermo Fisher Scientific), using the following pulse parameters: 1,350 V, 10 ms, and 3 pulses. The transfectants were split into two fractions 24 h after electroporation: one fraction was stimulated with PMA (40 ng/mL) and ionomycin (0.2 μM) for 24 h and the other was left unstimulated. Cells were harvested, and total RNA was extracted with TRIzol (Thermo Fisher Scientific) and the Monarch Spin RNA Isolation Kit (New England Biolabs) according to the manufacturers’ instructions. Total RNA (1 µg) was reverse transcribed to generate cDNA with the SuperScript™ IV First-Strand Synthesis System (Thermo Fisher Scientific). Quantitative PCR for IL17A was performed on a StepOnePlus™ Real-Time PCR System (Thermo Fisher Scientific). Transcript levels were determined in triplicate and normalized against GAPDH levels. Data were analyzed by the comparative threshold cycle (2^−ΔΔCt) method.

### High-throughput sequencing of *TRA/TRD*

Genomic DNA (gDNA) was extracted from peripheral blood obtained from healthy controls, age-matched controls, *RORC* WT/M family members, and *RORC* M/M patients P8–P12. The complementarity-determining region 3 (CDR3) of T-cell receptor alpha/delta (*TRA/TRD*) was amplified by multiplex PCR from gDNA templates (Adaptive Biotechnologies/ImmunoSEQ, Seattle, WA and CD genomics, Shirley, NY) as previously described (*50-53*). The Adaptive Biotechnologies protocol targeted both the *TRA* and *TRD* regions, whereas the CD genomics protocol was specific for *TRA*. Amplicons were sequenced with the Illumina NGS platform. Raw sequence data were processed with MiXCR, filtered for errors, and aligned with the reference genome based on the TCRα/δ (Bolotin et al., 2015), with V, D and J genes defined according to the reference sequence in the IMGT database (www.imgt.org). Sequences were retained for downstream analysis if they were productive, had a CDR3 amino-acid (AA) length of at least six residues, including a minimum of two sequence reads, and were of sufficient quality for V and J gene assignment. Clones with unresolved or ambiguously annotated V or J genes were excluded. Only analyses focusing on TCRα were performed. Clones containing *TRDV*, *TRDD*, and *TRDJ* sequences were removed. After these filtering steps, clone fractions were calculated by dividing the number of reads for each clone by the total number of qualified counts for each sample. Clonotype was defined by the combination of the *TRAV* gene, the *TRAJ* gene, and the corresponding CDR3 amino-acid sequence. Visualizations, including V-J usage heatmaps, circle-packing plots, Shannon entropy, diversity metrics (D50), and bubble heatmaps, were generated with the R packages *Immunarch* (https://github.com/immunomind/immunarch) and *ggplot2*.

### *In vitro* CD3^+^TCRαβ^+^ CD4^+^ T-cell polarization

Cryopreserved PBMCs were labeled with antibodies directed against CD45RA (BV605, clone HI100, #562886, BD Horizon), CCR7 (AF700, clone 150503, #561143, BD Pharmingen), CD3 (BV421, clone UCHT1, #562426, BD Horizon), CD8 (BUV395, clone RPA-T8, # 563795, BD Horizon), CD4 (APC, clone RPAT4, #300514, BioLegend), TCRαβ (PECy7, clone IP26, #306720, BioLegend), and TCRγδ (PE, clone B1.1, #12-9959-42, eBioscience), and naive (defined as CD45RA^+^CCR7^+^) and memory (defined as CD45RA^−^CCR7^+/-^) CD3^+^TCRαβ^+^CD4^+^ T cells were isolated (>98% purity) with a FACS Aria cell sorter (BD Biosciences). Isolated naive CD4^+^ T cells were then cultured with T-cell activation and expansion beads (TAE, anti-CD2/CD3/CD28, Miltenyi Biotec) + IL-2 (50 IU/mL, #IL002, Millipore) for 7 days to allow proliferation to occur. The cells were then harvested and recultured with TAE beads alone (TH0) or under TH1 (IL-12 [50 ng/mL, # 219-IL-005, R&D Systems]), TH2 (IL-4 [1 IU/mL, # 200-04, Peprotech], TH9 (IL-4 [100 IU/mL, # 200-04, Peprotech], TGF-β1 [2.5 ng/mL, #100-21C-10, Peprotech]), TH17 (TGF-β1 [2.5 ng/mL, #100-21C-10, Peprotech], IL-1β [50 ng/mL,#200-01B-10, Peprotech], IL-6 [50 ng/mL, #200-06-20, PeproTech], IL-21 [50 ng/mL, #200-21, PeproTech], or IL-23 [50 ng/mL, #200-23-10, PeproTech]) polarizing conditions. Isolated memory CD4^+^ T cells were cultured with TAE beads alone (TH0) or under TH1 or TH17 polarizing conditions. After five days of polarization, the supernatant was used for assessments of the secretion of IL-2, IL-4, IL-5, IL-6, IL-9, IL-10, IL-13, IL-17A, IL-17F, IFN-γ, and TNF with a cytometric bead array (BD Biosciences). Cytokine levels from naive CD4^+^ T cells were compared in Wilcoxon rank-sum tests with Benjamini-Hochberg (BH) correction. The cytokine levels of memory CD4^+^ T cells were compared in Wilcoxon rank-sum tests.

### PBMC BCG stimulation assay

Freshly thawed PBMCs were dispensed into a U-bottomed 96-well plate at a density of 2 × 10^5^ cells/well in 200 μL/well lymphocyte medium. Cells were incubated in the presence or absence of live BCG at a multiplicity of infection of 1, with or without recombinant human IL-12 (R&D Systems, 500 pg/mL), recombinant human IL-23 (Cat: 1290-IL; R&D Systems, 10 ng/mL), and IFN-γ1b (Actimmune, Amgen, 5000 U/mL). After 40 h of stimulation, GolgiPlug (Cat: 555029; BD Biosciences, 1:1,000) was added to each well to inhibit cytokine secretion. After another 8 h of incubation, supernatants were collected and stored at −20°C for the determination of cytokine levels, and the cells were harvested by centrifugation for flow cytometry analysis. The cells were stimulated simultaneously with PMA (Sigma-Aldrich, 25 ng/mL) and ionomycin (Sigma-Aldrich, 500 nM) for 8 h without GolgiPlug (for the determination of secreted cytokines) or for 1 h without GolgiPlug followed by 7 h with GolgiPlug (for intracellular cytokine staining) as a positive control. The levels of secreted cytokines in the supernatants were determined with the LEGENDplex Human Inflammation Panel 1 13-plex kit (Cat: 740809; BioLegend) and Th cytokine kit (Cat: 741027, BioLegend). Data were analyzed in R.

For flow cytometry analysis, cells were first stained with Zombie NIR dye (BioLegend) in PBS at room temperature for 15 min, then incubated on ice for 30 min with a surface-staining panel containing FcR blocking reagent (Miltenyi Biotec), 5-OP-RU-loaded MR1 tetramer-BV421 (NIH Tetramer Core Facility), anti-CD3-V450 (Clone UCHT1; BD Biosciences), anti-CD4-BUV563 (Clone SK3; BD Biosciences), anti-CD8-BUV737 (Clone SK1; BD Biosciences), anti-Vδ2-BUV805 (Clone B6; BD Biosciences), anti-CD123 (IL-3R)-BUV496 (BD Biosciences), anti-CD19-SparkNIR685 (Clone HIB19; BioLegend), anti-CD45-Alexa Fluor 532 (Thermo Fisher Scientific), anti-Vδ1-PerCP/Vio700 (Clone REA173; Miltenyi Biotec), anti-CD11c-BV750 (BD Biosciences), anti-γδTCR-BUV661 (Clone B1; BD Biosciences), anti-CD14-APC/Fire810 (Clone M5E2; BioLegend), anti-HLA-DR-Qdot655 (Invitrogen), anti-CD56-BV605 (Clone 5.1H11; BD Biosciences), anti-iNKT-BV480 (Clone 6B11; BD Biosciences), anti-Vβ11-APC-Vio770 (Clone REA559; Miltenyi Biotec), and anti-Vα7.2-PerCP/Cy5.5 (Clone 3C10; BioLegend) antibodies. Cells were fixed by incubation with 2% paraformaldehyde in PBS on ice for 15 min, then permeabilized and stained overnight at −20 °C in the perm buffer from the True-Nuclear Transcription Factor Buffer Set (BioLegend) with an intracellular cytokine panel containing FcR blocking reagent (Miltenyi Biotec), anti-T-bet-PE-Cy7 (Clone 4B10; BioLegend), anti-IFN-γ-BV711 (Clone 4S.B3; BioLegend), anti-IL-10-PE-Dazzle594 (Clone JES3-9D7; BioLegend), anti-TNF-BV510 (Clone MAb11; BioLegend), anti-IL-17A-BV785 (Clone BL168; BioLegend), and anti-RORγT-PE (Clone Q21-559; BD Biosciences) antibodies. Cells were acquired with an Aurora cytometer (Cytek). Data were manually gated with FlowJo and then imported into R for further analysis. Cellular composition was visualized by UMAP based on the expression levels for CD3, CD4, CD8, CD20, CD56, γδTCR, Vδ1, Vδ2, Vα7.2, MR1, T-bet, and RORγT, with the data downsampled to 10,000 cells per sample.

### Deep immunophenotyping

Freshly thawed PBMCs (~1.0-1.5 × 10^6^ cells) were simultaneously stained with LIVE/DEAD Fixable Blue (Cat: L23105; 1:800 in PBS; Invitrogen) and blocked by incubation with FcR Blocking Reagent (1:25; Miltenyi Biotec) on ice for 15 min. The cells were washed and surface-stained with the following reagents on ice for 30 min: Brilliant Stain Buffer Plus (Cat: 566385, 1:5; BD Biosciences), anti-γδTCR-BUV661 (Cat: 750019, Clone: 11F2, 1:50; BD Biosciences), anti-CXCR3-BV750 (Cat: 746895, Clone: 1C6, 1:20; BD Biosciences), and anti-CCR4-BUV615 (Cat: 613000, Clone: 1G1, 1:20; BD Biosciences) antibodies. Cells were then washed and surface-stained with the following reagents on ice for 30 min: anti-CD141-BB515 (Cat: 565084, Clone: 1A4, 1:40; BD Biosciences), anti-CD57-FITC (Cat: 347393, Clone: HNK-1, 3:250; BD Biosciences), anti-Vδ2-PerCP (Cat: 331410, Clone: B6, 3:500; BioLegend), anti-Vα7.2-PerCP-Cy5.5 (Cat: 351710, Clone: 3C10, 1:40; BioLegend), anti-Vδ1-PerCP-Vio700 (Cat: 130-120-441, Clone: REA173, 1:100; Miltenyi Biotec), anti-CD14-Spark Blue 550 (Cat: 367148, Clone: 63D3, 1:40; BioLegend), anti-CD1c-Alexa Fluor 647 (Cat: 331510, Clone: L161, 1:50; BioLegend), anti-CD38-APC-Fire 810 (Cat: 356644, Clone: HB-7, 3:100; BioLegend), anti-CD27-APC H7 (Cat: 560222, Clone: M-T271, 1:50; BD Biosciences), anti-CD127-APC-R700 (Cat: 565185, Clone: HIL-7R-M21, 1:50; BD Biosciences), anti-CD19 Spark NIR 685 (Cat: 302270, Clone: HIB19, 3:250; BioLegend), anti-CD45RA-BUV395 (Cat: 740315, Clone: 5H9, 3:250; BD Biosciences), anti-CD16-BUV496 (Cat: 612944, Clone: 3G8, 3:500; BD Biosciences), anti-CD11b-BUV563 (Cat: 741357, Clone: ICRF44, 1:100; BD Biosciences), anti-CD56-BUV737 (Cat: 612767, Clone: NCAM16.2, 3:250; BD Biosciences), anti-CD4-cFluor 568 (Clone: SK3, 3:250; Cytek), anti-CD8-BUV805 (Cat: 612889, Clone: SK1, 3:250; BD Biosciences) antibodies, MR1 tetramer-BV421 (1:100; National Institutes of Health Tetramer Core Facility), anti-CD11c-BV480 (Cat: 566135, Clone: B-ly6, 1:40; BD Biosciences), anti-CD45-BV510 (Cat: 563204, Clone: HI30, 3:250; BD Biosciences), anti-CD33-BV570 (Cat: 303417, Clone: WM53, 3:250; BioLegend), anti-iNKT-BV605 (Cat: 743999, Clone: 6B11, 1:25; BD Biosciences), anti-CD161-BV650 (Cat: 563864, Clone: DX12, 1:25; BD Biosciences), anti-CCR6-BV711 (Cat: 353436, Clone: G034E3, 3:250; BioLegend), anti-CCR7-BV785 (Cat: 353230, Clone: G043H7, 1:40; BioLegend), anti-CD3-Pacific Blue (Cat: 344824, Clone: SK7, 3:250; BioLegend), anti-CD20-Pacific Orange (Cat: MHCD2030, Clone: HI47, 1:50; Invitrogen), anti-CD123-Super Bright 436 (Cat: 62-1239-42, Clone: 6H6, 1:40; Invitrogen), anti-Vβ11-PE (Cat: 130-123-561, Clone: REA559, 3:500; Miltenyi Biotec), anti-CD24-PE-Alexa Fluor 610 (Cat: MHCD2422, Clone: SN3, 1:25; Invitrogen), anti-CD25-PE-Alexa Fluor 700 (Cat: MHCD2524, Clone: 3G10, 1:25; Invitrogen), anti-CRTH2-biotin (Cat: 13-2949-82, Clone: BM16, 1:50; Invitrogen), anti-CD209-PE-Cy7 (Cat: 330114, Clone: 9E9A8, 1:25; BioLegend), anti-CD117-PE-Dazzle 594 (Cat: 313226, Clone: 104D2, 3:250; BioLegend), anti-HLA-DR-PE-Fire 810 (Clone: L243, 1:50; BioLegend), and anti-CD66b-APC (Cat: 1305118, Clone: G10F5 1:50; eBioscience) antibodies. After washing, cells were further incubated with streptavidin-PE-Cy5 (Cat: 405205, 1:3,000; BioLegend) on ice for 30 min. Cells were then washed, fixed with 1% paraformaldehyde in PBS, washed again, and acquired on an Aurora cytometer (Cytek). Subsets were manually gated with FlowJo, and results were visualized with R.

### scRNA-seq

We performed scRNA-seq analyses on cryopreserved PBMCs from six healthy adult controls, four RORγ/ RORγT-deficient patients, and three IL-12Rβ1-deficient patients. Cells were filtered through a MACS SmartStrainer with 70 µm pores (Cat: 130-098-462; Miltenyi Biotec) to remove large debris, washed three times with PBS plus 0.5% FBS, and filtered through a Falcon Cell Strainer with 40 µm pores (Cat: 352340; Corning) before being subjected to single-cell capture via the 10X Genomics Chromium chip. Libraries were prepared with the Chromium Single Cell 3′ Reagent Kit (v3 Chemistry) and sequenced with an Illumina NovaSeq 6000 sequencer. Sequences were preprocessed with CellRanger. Approximately 10,000 cells were sequenced per sample. For the baseline scRNA-seq analysis, the data generated during this study were analyzed together with the data for healthy controls generated in the previous study (Ogishi et al., 2023a), and publicly available control PBMC datasets downloaded from the 10X Genomics web portal (https://support.10xgenomics.com/single-cell-gene-expression/datasets). Data were filtered manually based on common quality-control metrics and integrated with Harmony (Korsunsky et al., 2019). Two sequential graph-based clustering analyses were performed. The first-round clustering identified general leukocyte subsets, and the second-round clustering identified memory and effector T-lymphocyte subsets and NK lymphocytes at sufficiently high resolution. Clusters were identified with the SingleR pipeline guided by MonacoImmuneData (Aran et al., 2019), and cell type-specific marker gene expression was then assessed by manual inspection (Monaco et al., 2019). Clusters were visualized by Uniform Manifold Approximation and Projection (UMAP). After cluster identification, 10X datasets were excluded from subsequent analyses. Gene expression was quantified at single-cell level with Seurat (Hao et al., 2021). Pseudobulk analysis was performed by aggregating all reads from cells assigned to a given cluster, as previously described (Crowell et al., 2020). We performed PCA on the read counts normalized through variance-stabilizing transformation with batch correction, using the removeBatchEffect function implemented in limma (Ritchie et al., 2015). Differential expression analysis was performed with DESeq2 (Love et al., 2014). WGCNA was performed in R (Langfelder and Horvath, 2008). TFEA was performed with ChEA3 (https://maayanlab.cloud/chea3/) (Keenan et al., 2019). All analyses were performed in R v4 (http://www.R-project.org/). Raw data are available from the SRA BioProject, accession no. PRJNA856671. For the baseline scRNA-seq analysis, data generated during this study were analyzed together with the data for healthy controls and control patients generated in the previous study (Arias et al., 2024; Johnson et al., 2024; Philippot et al., 2023). Immunity-focused pathway analysis was performed with the MSigDB C7 immunologic signatures collection/ImmuneSigDB (Godec et al., 2016; Liberzon et al., 2015; Subramanian et al., 2005). Focused module analysis was performed in naïve CD4^+^ and CD8^+^ T cells with manually curated immune/T-cell gene modules selected to represent the major biological programs observed in the unbiased analyses. The modules included naïveness (*IL7R, LTB, LEF1, TCF7, MAL, SELL, MALAT1, CCR7*), IL-2/STAT5 signaling (*IL7R, LTB, MAL, TCF7, LEF1, BCL2, PIM1, STAT5A*), TNF/NF-κB activation (*NFKBIA, TNFAIP3, JUNB, FOS, BCL3, RELB, DUSP1, CXCL2, CCL3, CCL4*), IL-6/STAT3 signaling (*STAT3, SOCS3, JUN, FOS, BCL3, ICAM1, NAMPT, PIM1*), type I IFN response (*ISG15, IFIT1, IFIT2, IFIT3, MX1, OAS1, IFI6, IFI44L, RSAD2, BST2*), type II IFN response (*STAT1, IRF1, GBP1, GBP5, CXCL10, TAP1, B2M, HLA-B, IFITM1, WARS*), TGF-β signaling (*SMAD3, TGFBR2, LTBP1, TAGLN, TSC22D1, KLF10, SERPINE1, ENG*), and cellular stress/UPR (*DDIT3, ATF3, JUN, FOS, HSPA5, XBP1, TXNIP, DNAJB9*). Only genes detected in the scRNA-seq object were used for scoring. Module scores were calculated with Seurat addModuleScore (Hao et al., 2021), summarized by donor and cell type, and compared between patients with RORγ/RORγT deficiency and controls in Wilcoxon rank-sum tests with false discovery rate correction. We analyzed TF-family activity by ranking pseudobulk differential-expression results and applying fgsea to MSigDB C3 transcription factor target/motif gene sets (Liberzon et al., 2015). Representative enriched TF-family signatures were retained for naïve CD4^+^ and CD8^+^ T cells and combined across naïve T-cell subsets by signed enrichment direction.

### Statistical analysis

Student’s *t*-tests, Mann-Whitney *U* tests, one-way ANOVA, and two-way ANOVA were used on the datasets, to investigate the significance of differences. In all the figures, bar graphs represent either the mean and standard deviation or the mean and standard error of the mean. Dots correspond to individual samples or technical replicates. *P* values of 0.05 and below are considered statistically significant. \**p*<0.05, \*\**p*<0.01, \*\*\**p*<0.001, \*\*\*\**p*<0.0001, and ns = not significant (or not marked). Detailed statistical methods and analyses are provided in the legends to the figures.

## Non-standard abbreviations

AR: autosomal recessive
BCG: Bacille Calmette-Guérin
BCG-osis: disseminated BCG vaccine infection
CMC: chronic mucocutaneous candidiasis
IEI: inborn error of immunity
IFN-γ: interferon gamma
ILC: innate lymphoid cell
iNKT: invariant natural killer T cell
LOF: loss-of-function
MAIT: mucosa-associated invariant T cell
MSMD: Mendelian susceptibility to mycobacterial disease
Mtb: *Mycobacterium tuberculosis*
PBMC: peripheral blood mononuclear cell
RORγT: retinoic acid-related orphan receptor gamma T
scRNA-seq: single-cell RNA sequencing
TB: tuberculosis
TCR: T-cell receptor
TH: T helper
TREC: T-cell receptor excision circle
WES: whole-exome sequencing
WT: wild-type

## Supplemental Material

Fig. S1 supplements Fig. 1 and Fig. 2 and shows that V45F/V24F, K381I/K460I, Y186*/Y165*, T421Nfs*56/ T400Nfs*56, and R98*/R77* *RORC* variants are LOF when overexpressed, whereas common population *RORC* variants are functionally neutral. Fig. S2 supplements Fig. 3 and shows intact TCRα rearrangement heterozygous carriers of LOF *RORC* variants, as well as the patterns of IFN-γ and IL-17A production by lymphocyte subsets upon stimulation. Fig. S3 supplements Fig. 4 and shows deep immunophenotyping and longitudinal T-cell trends in patients with AR RORγ/RORγT deficiency. Fig. S4 supplements Fig. 5 and shows memory/exhaustion-associated skewing and distinct transcriptomic profiles in naïve T cells from patients with AR RORγ/RORγT deficiency. Fig. S5 supplements Fig. 7 and shows IFN-γ and TNF production by various lymphocyte subsets during mycobacterial infection.

## DATA AVAILABILITY STATEMENT

The data are available from the corresponding author upon reasonable request, subject to the consent provisions of the patients involved.

## ACKNOWLEDGMENTS

We thank the patients and their families; the members of the laboratory for helpful discussions; Tatiana Kochetkov and Marianna Teplova for technical assistance; and Yelena Nemirovskaya, Dana Liu, Mark Woollett, Kerel Francis, Maya Chrabieh, and Lazaro Lorenzo-Diaz for administrative assistance. We thank physicians, including but not limited to Eguchi Katsuhide, Masataka Ishimura (Department of Pediatrics, Graduate School of Medical Sciences, Kyushu University, Fukuoka, Japan), Youta Kuroki, and Shigenobu Tokunaga (Division of Pediatrics, Faculty of Medicine, University of Miyazaki, Miyazaki, Japan), the patients, and their families. We thank the Genomics Resource Center and Flow Cytometry Resource Center (RRID:SCR_017694) for technical support and the Rockefeller University Hospital for patient-oriented support. We also thank the National Institutes of Health (NIH) Tetramer Core Facility (NTCF) for providing the MR1 tetramer, which was developed jointly with Dr. James McCluskey, Dr. Jamie Rossjohn, and Dr. David Fairlie.

## AUTHOR CONTRIBUTIONS

I.F. conceptualized the study, designed the experiments, interpreted data, and wrote the manuscript. M.T. and A.G. conducted experiments and analyzed data. H.A. referred patients, conducted experiments, and analyzed data. S.Sha. gathered clinical histories, collected blood samples for research, and referred patients. M.Mes. gathered clinical histories, collected blood samples for research, and referred patients. T.N. gathered clinical histories, collected blood samples for research, and referred patients. H.P.L. gathered clinical histories, collected blood samples for research, and referred patients. S.Rao gathered clinical histories, collected blood samples for research, and referred patients. St.Ri. gathered clinical histories, collected blood samples for research, and referred patients. J.E.H. conducted experiments and analyzed data. O.M.D. conducted experiments and analyzed data. C.K. conducted experiments and analyzed data. J.G.M. conducted experiments and analyzed data. M.O. conducted experiments and analyzed data. J.H. conducted experiments and analyzed data. J.P. conducted experiments and analyzed data. J.V. analyzed high-throughput sequencing data. Y.F. generated figures. C. Sou. conducted experiments and analyzed data. M. Mig. conducted experiments and analyzed data. B.P. provided analytical support. K.J.L.J. provided technical support and analytical advice. S. Nis. conducted experiments and analyzed data. So. S. conducted experiments and analyzed data. K.K. conducted experiments and analyzed data. A.Y. gathered clinical histories, collected blood samples for research, and referred patients. H.M. gathered clinical histories, collected blood samples for research, and referred patients. M.A. gathered clinical histories, collected blood samples for research, and referred patients. F.V. gathered clinical histories, collected blood samples for research, and referred patients. T.C. gathered clinical histories, collected blood samples for research, and referred patients. J. Sma. gathered clinical histories, collected blood samples for research, and referred patients. S.Ch. gathered clinical histories, collected blood samples for research, and referred patients. Z.C. gathered clinical histories, collected blood samples for research, and referred patients. Sh. A. gathered clinical histories, collected blood samples for research, and referred patients. A.T. conducted experiments and analyzed data. P.Z. analyzed high-throughput sequencing data. J.R. interpreted data and provided intellectual contributions to the study. L.D.N. interpreted data and provided intellectual contributions to the study. Q.P.-H. interpreted data and provided intellectual contributions to the study. S.G.T. and C.S.M. conceptualized and supervised the study, designed the experiments, interpreted data, analyzed data, generated figures, and edited the manuscript. J.-L.C. conceptualized and supervised the study, wrote and revised the manuscript, provided laboratory infrastructure, and secured funding. S.O. conceptualized the study, supervised the study, designed the experiments, conducted experiments, interpreted data, and edited the manuscript. A.P. supervised the study, designed the experiments, interpreted data, and edited the manuscript. J.B. supervised the study, designed the experiments, interpreted data, and edited the manuscript. S.B.-D. supervised the study, designed the experiments, conducted experiments, interpreted data, and edited the manuscript. R.Y. conceptualized the study, supervised the study, designed the experiments, conducted experiments, interpreted data, analyzed data, illustrated the figures, wrote, and revised the manuscript.

## FIGURE LEGENDS

**Figure S1.**
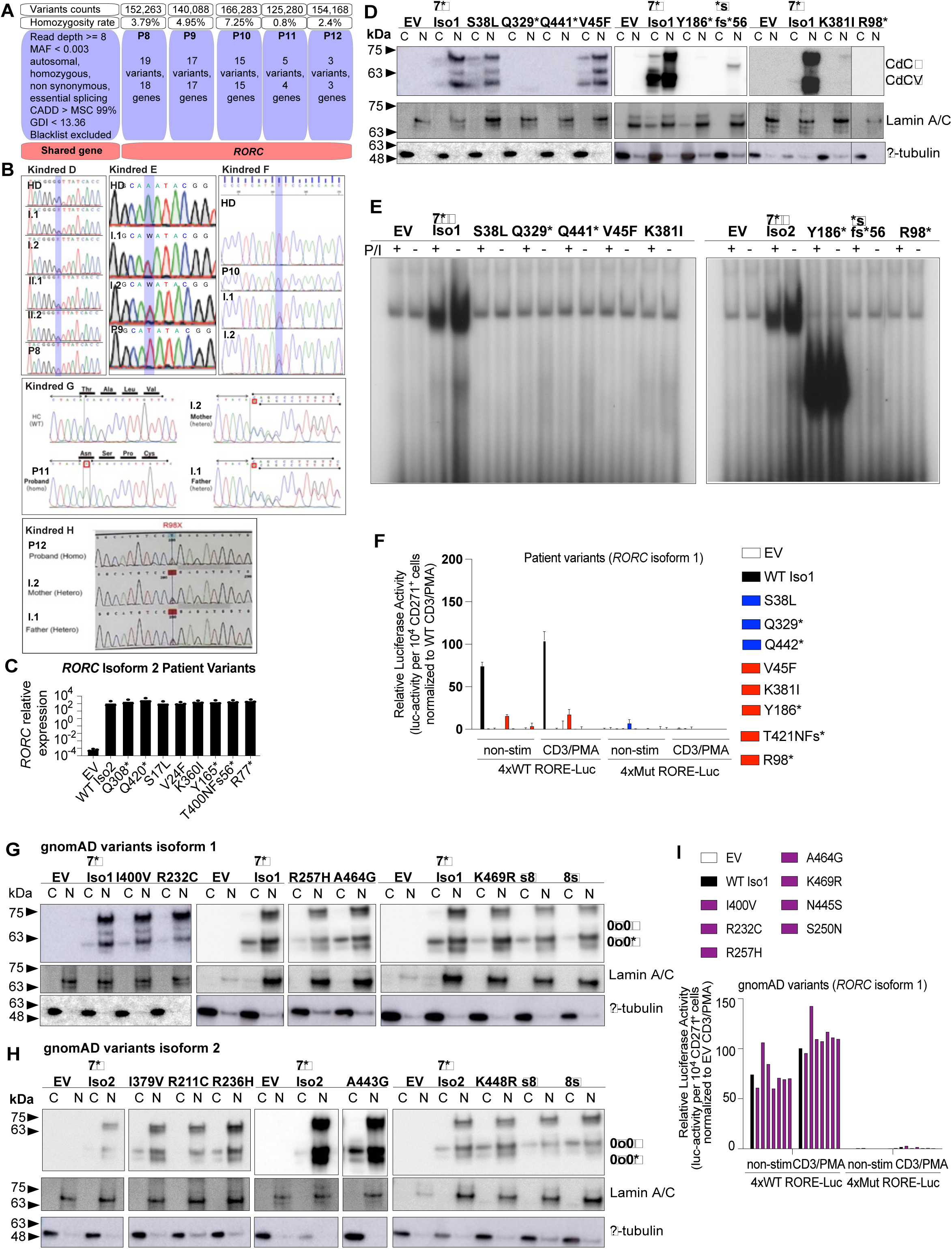
Genome-wide analyses of RORγ/RORγT-deficient patients and functional characterization of the mutant and populational *RORC* variants in an overexpression system. (**A**) Schematic diagram depicting the workflow for the identification of the five new rare variants in *RORC.* (**B**) Sanger sequencing results to confirm the genotypes of P8 – P12 harboring new homozygous *RORC* variants. (**C**) Relative expression of *RORC* measured by quantitative RT-PCR (RT-qPCR) following the transfection of HEK293T cells with WT and mutant pCMV6-*RORC*-Myc-DDK plasmids. (**D**) Western-blot analysis of WT or mutant *RORC* isoform 1 (RORγ) variants to assess protein levels and subcellular distribution in HEK293T cells. C, cytoplasmic fraction; N, nuclear fraction. (**E**) Electromobility shift assay (EMSA) with a ^32^P-labeled RORE-2 probe derived from the *IL17A* promoter, incubated with nuclear lysates of HEK239T cells transfected with the indicated vectors encoding RORγ, with or without PMA/ionomycin stimulation (P/I). (**F**) Transcriptional activity of patient RORγ variants in an *in vitro* luciferase reporter system driven by WT or mutant multimerized RORE. (**G** and **H**) Western-blot analysis of mutant *RORC* isoform 1 (RORγ) (G) and isoform 2 (RORγT) (H) variants from gnomAD to assess protein levels and subcellular distribution in HEK293T cells. C, cytoplasmic fraction; N, nuclear fraction. (**I**) Transcriptional activity of gnomAD variants shown in (G), as determined with an *in vitro* luciferase reporter system driven by WT or mutant multimerized RORE. All experimental data were verified in at least 2 independent experiments.

**Figure S2.**
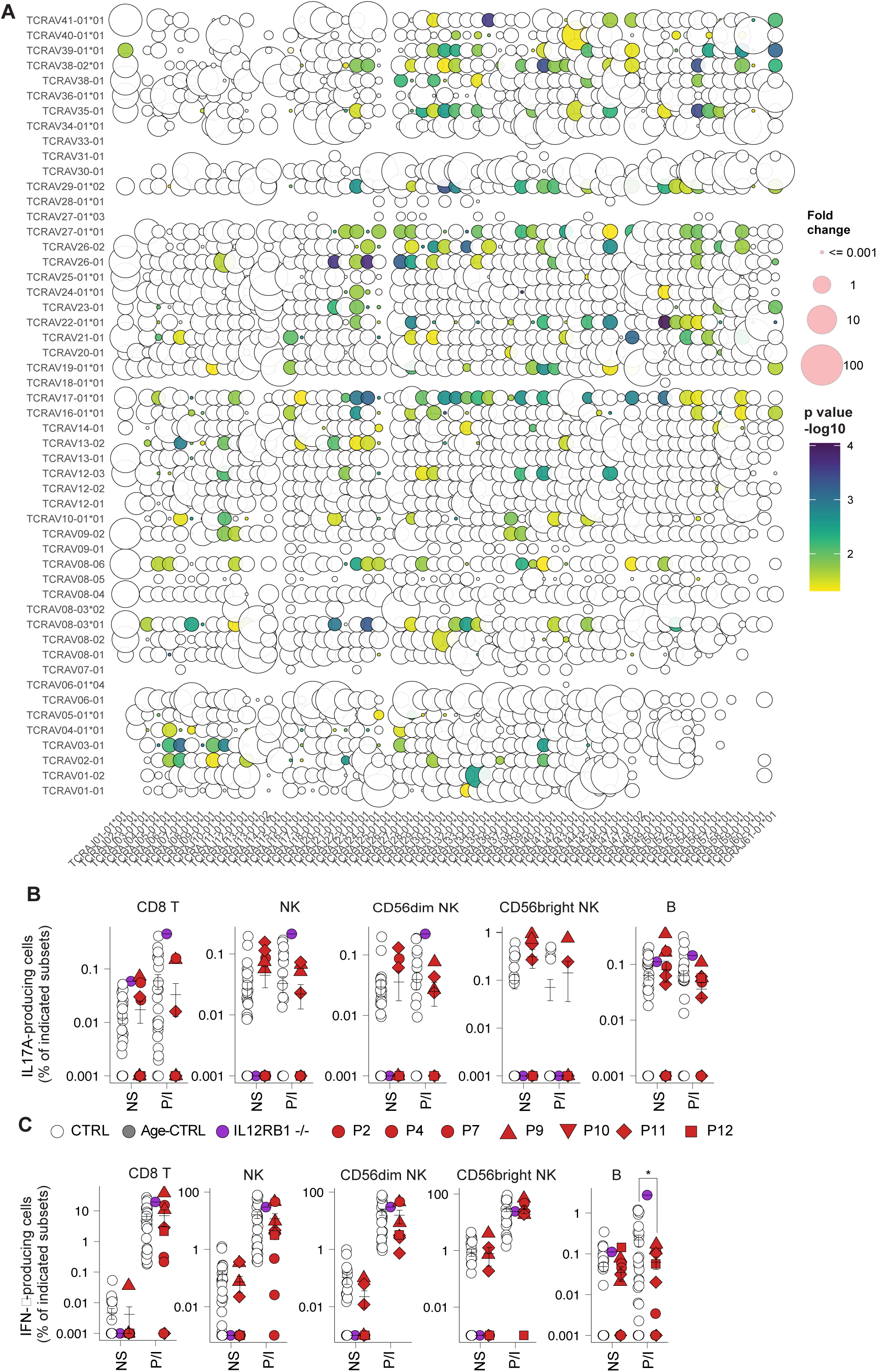
Normal TCRα rearrangement in heterozygous parents and similar levels of IL-17A production between patients and controls for certain lymphocyte subsets. (**A**) Bubble plot of high-throughput sequencing of the *TRAV/TRAJ* loci in genomic DNA from healthy controls and seven WT/M heterozygous carriers of loss-of-function *RORC* variants. Fold-change difference in *TRAV* and *TRAJ* usage in WT/M carriers relative to controls is indicated by bubble size, and statistical significance (*p*-value) by color, with *TRAV* segments on the *y*-axis and *TRAJ* segments on the *x*-axis. (**B and C**) Frequency of IL-17A-producing (B) and IFN-γ-producing cells (C) within the indicated subsets in controls or patients, with or without P/I stimulation. All experimental data were verified in at least 2 independent experiments.

**Figure S3.**
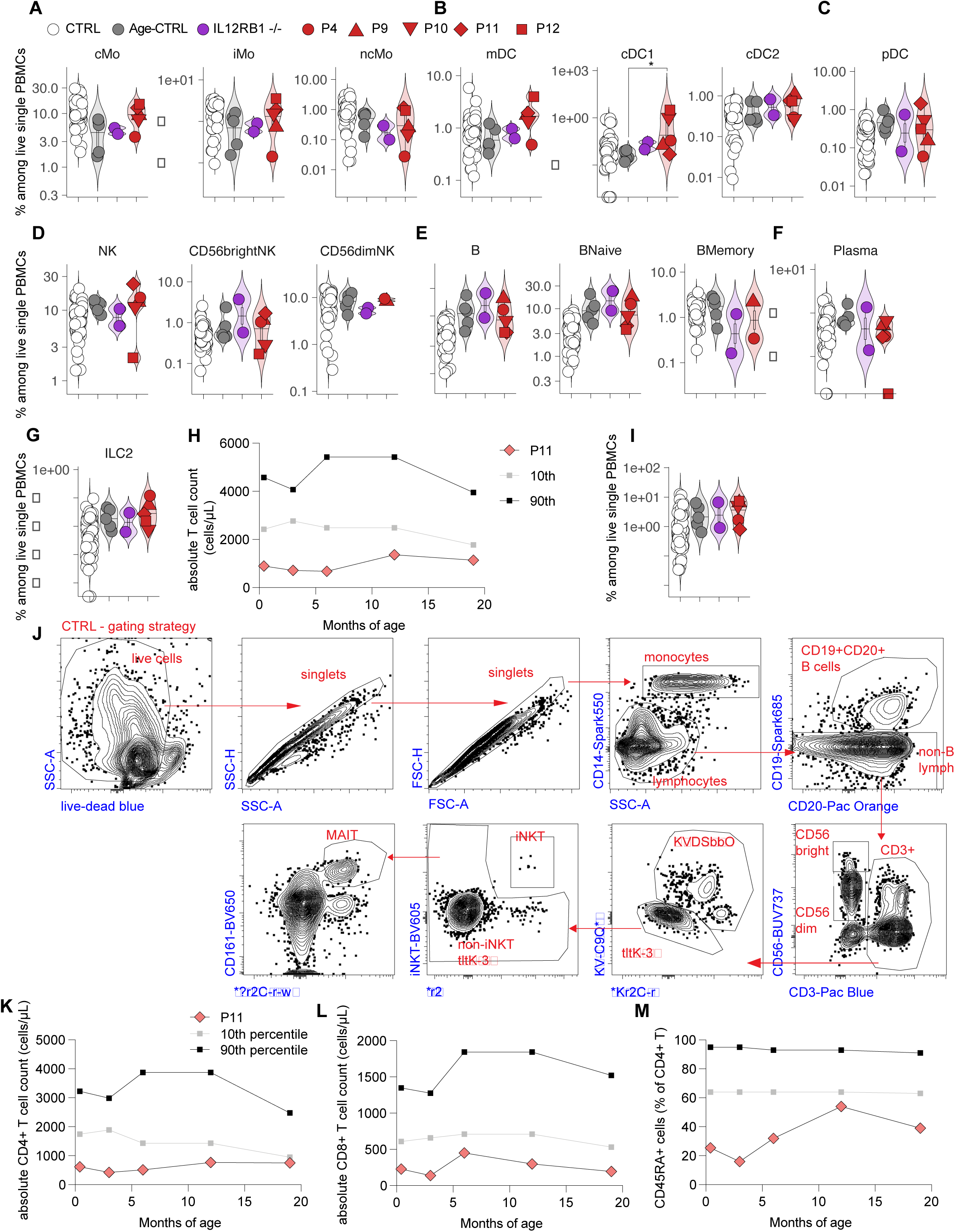
Deep immunophenotyping and trends over time in patients with AR RORγ/RORγT deficiency. (**A - G**) Frequencies of monocyte subsets (A), myeloid dendritic cell subsets (B), plasmacytoid dendritic cells (C), NK cell subsets (D), B-cell subsets (E), plasma cells (F), and ILC2 cells (G), among live PBMCs from controls, age-matched controls, and patients with AR IL-12Rβ1 or RORγ/RORγT deficiencies. (**H**) Longitudinal trend in the absolute T-cell counts of P11 relative to the reference range for age. (**I**) Frequencies of Vδ2^+^ γδ T cells among live PBMCs from controls, age-matched controls, and patients with AR IL-12Rβ1 or RORγ/RORγT deficiencies. (**J**) Gating strategy used for the identification of MAIT and iNKT cells. (**K - M**) Longitudinal trends in absolute CD4^+^ T-cell counts (K), absolute CD8^+^ T-cell counts (L), and frequencies of CD45RA^+^ CD4^+^ T cells (M) in P11 relative to the reference ranges for age. All experimental data were verified in at least 2 independent experiments.

**Figure S4.**
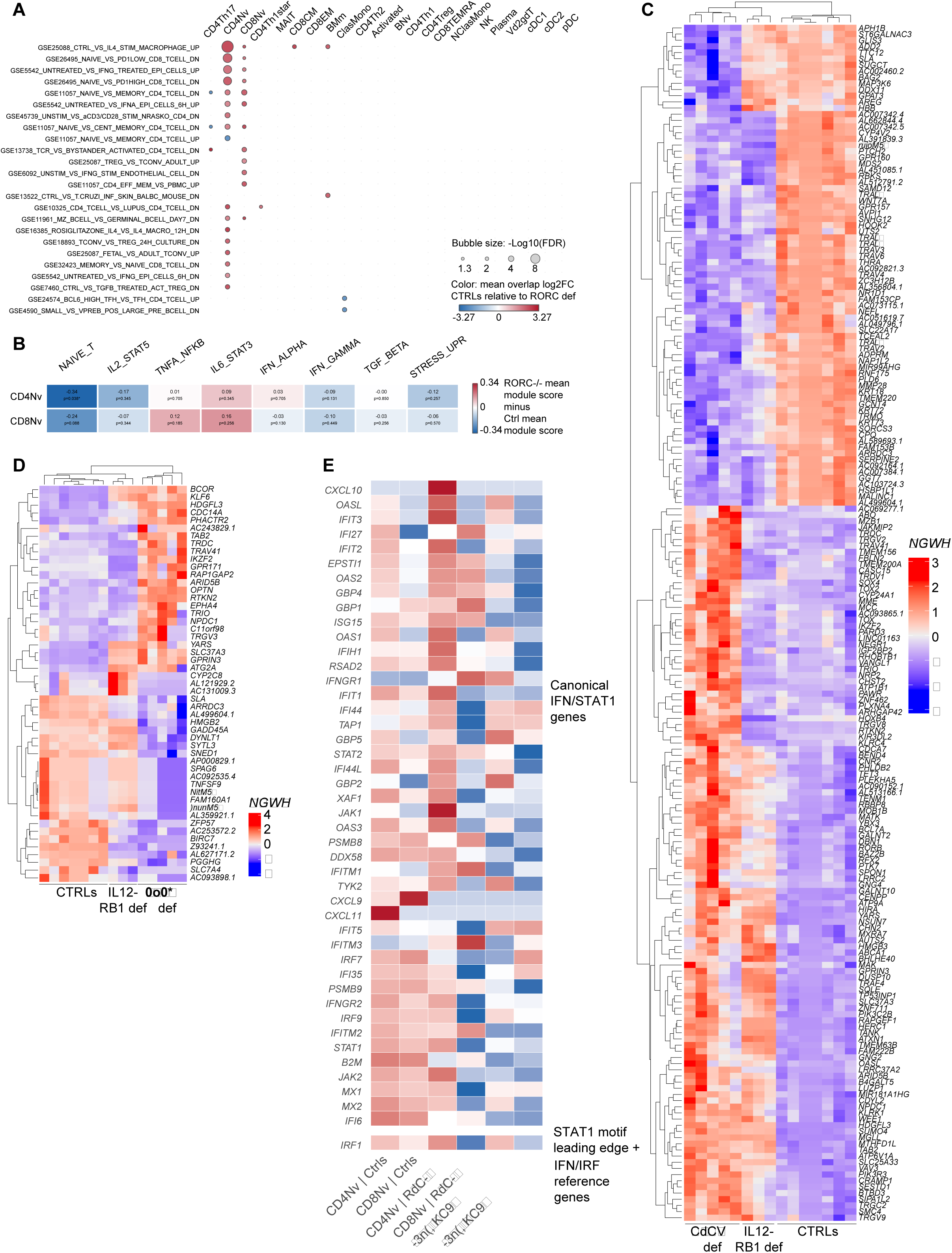
Skewing toward memory and exhaustion-associated phenotypes and distinct transcriptomic profiles of naïve T cells from patients with AR RORγ/RORγT deficiency. (**A**) ImmuneSigDB C7 enrichment across pseudobulked immune subsets in RORγ/RORγT deficiency versus controls. Color indicates fold-change direction; size indicates −log10 false-discovery rate. (**B**) Focused module scores in naïve CD4^+^ and CD8^+^ T cells. Color indicates mean score difference between RORγ/RORγT deficiency and controls. All experimental data were verified in at least 2 independent experiments. (**C and D**) Heatmap of differentially regulated genes in naïve CD4^+^ T cells (C) and CD8^+^ (D) T cells across controls, IL-12Rβ1 deficiency, and RORγ/RORγT deficiency. (**E**) Canonical IFN/STAT1 gene-expression heatmap in naïve CD4^+^ and CD8^+^ T cells, used to assess whether STAT1-associated TF enrichment reflects type I or type II interferon biology.

**Figure S5.**
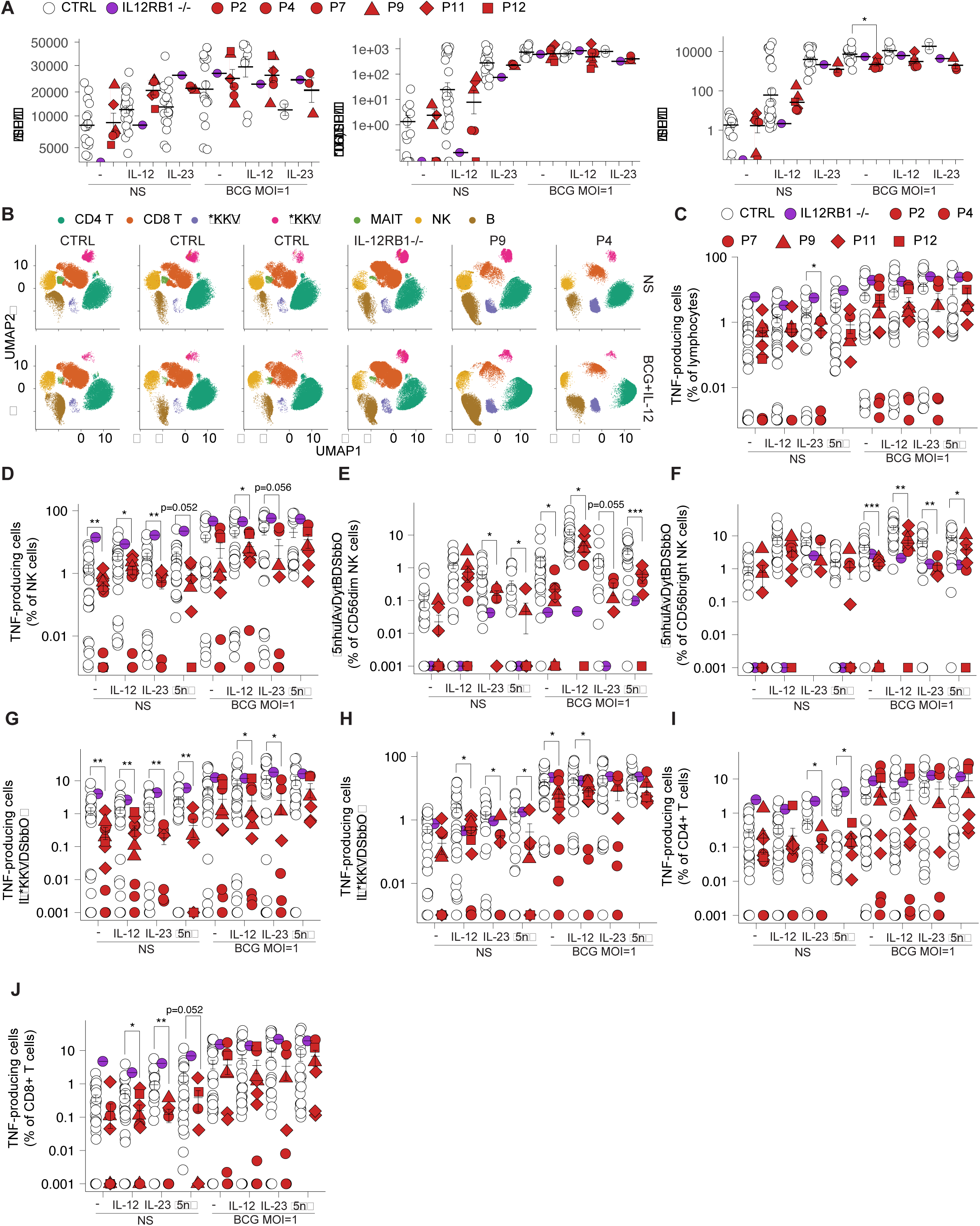
Analysis of IFN-γ and TNF production during mycobacterial infection. (**A**) Production of IL-8, GM-CSF, and IL-1β by PBMCs with and without live BCG infection, in the presence or absence of exogenous IL-12 or IL-23. (**B**) UMAP visualization of leukocyte subsets in PBMCs from healthy controls, patients with IL-12Rβ1 deficiency, and patients with RORγ/RORγT deficiency. (**C and D**) Frequencies of TNF-producing cells among total lymphocytes (C), and among NK cells (D), following *in vitro* infection with live BCG, in the presence or absence of exogenous IL-12, IL-23, or IFN-γ. (**E and F**) Frequencies of IFN-γ-producing cells among CD56^dim^ NK cells (E) and CD56^bright^ NK cells (F), following *in vitro* infection with live BCG, in the presence or absence of exogenous IL-12, IL-23, or IFN-γ. (**G - J**) Frequencies of TNF-producing cells among Vδ1^+^ γδ T cells (G), Vδ2^+^ γδ T cells (H), CD4^+^ T cells (I), and CD8^+^ T cells (J), following *in vitro* infection with live BCG, in the presence or absence of exogenous IL-12, IL-23, or IFN-γ. All experimental data were verified in at least 2 independent experiments.

